# Safety and tolerability of electronic cigarettes to reduce cigarette smoking: Secondary analysis from a randomized placebo-controlled trial

**DOI:** 10.64898/2026.03.18.26348637

**Authors:** Sitasnu Dahal, Soha Talih, Shari Hrabovsky, Christopher Sciamanna, Craig Livelsberger, Eric Soule, Caroline O. Cobb, Jessica Yingst, Jonathan Foulds

**Author notes:** Corresponding Author: Sitasnu Dahal, 500 University Drive, Hershey, PA 17033,.

## Abstract

**Background:** The clinical safety profile of e-cigarette use for smoking reduction remains poorly characterized. This study compared the relative safety and tolerability of nicotine e-cigarette use with non-nicotine e-cigarettes or a non-aerosol cigarette substitute (CS) among adults interested in reducing their smoking.

**Methods:** We conducted a secondary analysis of adverse events (AEs) reported in a 6-month, double-blind RCT involving 520 participants assigned to either e-cigarettes with 0, 8, or 36 mg/mL nicotine or a CS. AEs were coded using CTCAE V4.0 and assessed for frequency, severity, seriousness and relatedness across groups. Cumulative incidence was calculated over 24 weeks. We estimated risk differences (RDs) and 95% confidence intervals (CIs) for frequently reported AEs (≥1% of participants overall) comparing e-cigarette vs. CS and nicotine versus non-nicotine e-cigarette groups. Fisher’s exact test, with adjustment for multiple comparisons, was used to assess statistical significance.

**Results:** Most study-related AEs (those rated as possibly, probably, or definitely related by medical monitor) were mild in severity and none were classified as serious. At 24 weeks, cumulative incidence of first study-related AE was highest in the 36 mg/mL (37.0%) and 8 mg/mL (35.2%) e-cigarette groups, followed by 0 mg/mL (23.4%), and lowest in CS group (2.5%). E-cigarette users experienced significantly greater risks of cough (RD [95%CI]: 8.5% [5.6 – 11.3]), headache (RD [95%CI]: 5.4% [3.3 – 7.6]) and sore throat (RD [95%CI]: 5.4% [3.2 – 7.6]) as compared with the CS group. Cough was also more common in those randomized to nicotine versus non-nicotine e-cigarettes (RD [95%CI]: 8.1% [3.4 – 12.8]).

**Conclusion:** All study products were generally well-tolerated; however, AEs were more common in e-cigarette groups, especially with nicotine. Findings highlight the need to monitor common symptoms such as cough, headache, and sore throat in clinical and regulatory evaluations of e-cigarette safety.

**What is already known on this topic:** Nicotine e-cigarettes can help people who smoke to quit combustible cigarette use and reduce some cigarette-related toxicant exposures. However, the safety of inhaled nicotine and other constituents such as propylene glycol, vegetable glycerin in e-cigarettes remains unclear.

**What this study adds:** This study suggests that e-cigarette use is associated with a higher incidence of study-related AEs such as cough, headache, and sore throat, particularly among those using nicotine-containing products. However, overall safety and tolerability profiles were comparable across e-cigarette groups with differing nicotine concentrations.

**How this study might affect research, practice or policy:** These findings offer methodological guidance for evaluating e-cigarette safety in clinical trials and may inform regulators, clinicians, and public health professionals regarding the tolerability of e-cigarette products varying in nicotine concentration.

## INTRODUCTION

Smoking remains a leading cause of preventable morbidity and mortality worldwide, affecting nearly every organ and increasing the risk of lung cancer, cardiovascular disease, respiratory disease, and numerous other conditions.^1,2^ Despite well-established health risks, many individuals who smoke struggle to quit, primarily due to nicotine dependence.^3^ First-line interventions such as behavioral counseling and U.S. Food and Drug Administration (FDA)-approved pharmacotherapies like nicotine replacement therapies (NRTs), bupropion, and varenicline have proven efficacy but are not acceptable for all users.^4–6^ As a result, many people who smoke turn to alternative nicotine delivery systems.^7^ One such product that has gained popularity over nearly two decades is electronic cigarettes (e-cigarettes).

E-cigarettes are devices that work by heating a liquid containing primarily nicotine, propylene glycol (PG), vegetable glycerin (VG), and flavoring agents to produce an aerosol that is inhaled via a mouthpiece.^8^ E-cigarettes may appeal to some because they can deliver nicotine, are available in many flavors, and using e-cigarettes resembles behavioral aspects of smoking.^9^ Clinical trials and systematic reviews indicate that when used by people who smoke combustible cigarettes, nicotine containing e-cigarettes can reduce exposure to toxicants (e.g., 4-(methylnitrosamino)-1-(3-pyridyl)-1-butanol [NNAL] and exhaled carbon monoxide [CO]) and may increase smoking cessation rates compared to placebo e-cigarettes or NRTs.^10–12^ However, the overall risk-benefit profile of e-cigarette use, particularly among individuals who continue to smoke, remains incompletely understood due to a lack of long-term data from controlled clinical trials.^13^

While e-cigarettes are generally considered less harmful than combustible cigarettes for people who smoke,^9^ concerns persist regarding the adverse health effects of inhaling aerosolized constituents such as PG, VG, and nicotine, especially at high concentrations or with long-term use.^14^ A major limitation in the current evidence base, highlighted by systematic reviews and meta-analyses, including Cochrane living systematic review for smoking cessation, is the scarcity of robust data on adverse events (AEs), serious adverse events (SAEs), and other safety indicators.^10,15–17^ Existing studies often provide low- to very-low-certainty evidence, with wide confidence intervals and inconsistent reporting, making conclusions tentative and subject to change as more data emerge.^10,16–19^ The current evidence emphasizes the need for improved, standardized safety monitoring in clinical trials to allow meaningful comparison of AE profiles across products, doses, and study designs.

This gap in safety data also has regulatory implications. In the United States, manufacturers seeking FDA marketing authorization for e-cigarette products must demonstrate that their products are “appropriate for the protection of public health,” which includes evidence of both effects on reducing cigarette smoking and without increasing serious AEs or recruiting a large number of non-smokers.^20^ Emerging evidence suggests that even mild AEs such as mouth/throat irritation, nausea, dizziness, nicotine dependence may reduce adherence to e-cigarettes and approved pharmacotherapies like NRTs,^21–24^ and can impede quitting success.^25^ As newer generations of e-cigarettes with higher nicotine concentrations, that closely mimic cigarette-like nicotine delivery, and flavors other than tobacco enter the market, including those authorized through the FDA’s premarket tobacco product application (PMTA) pathway, it is important to assess their safety profile.^26^ In particular, understanding the frequency and nature of AEs, especially those likely attributable to inhalation of PG, VG, and nicotine, is critical.

To address these gaps, we conducted a secondary analysis from a four-arm, double-blind randomized controlled trial (RCT) that was originally designed to examine the effects of e-cigarette use on tobacco-related toxicant exposure among individuals attempting to reduce their combustible cigarette use.^11^ This prespecified analysis aimed to characterize the frequency, severity, seriousness, and relatedness of AEs across e-cigarettes of varying nicotine concentrations (0, 8, and 36 mg/mL nicotine) and a non-aerosol, non-nicotine cigarette substitute group. We specifically evaluated study-related AEs, those deemed possibly, probably or definitely related to either the assigned study product or the study procedures, given their relevance to safety evaluations and regulatory decisions. Based on the known irritant/inflammatory properties of PG, VG, and nicotine,^27,28^ we hypothesized that study-related AEs would be more common among participants assigned to e-cigarette groups compared to the cigarette substitute group, and more frequent among nicotine-containing e-cigarettes than non-nicotine e-cigarette groups.

## METHODS

### Study design

This was a two-site, 9-month (6 months intervention and 3 months follow-up period), four-arm, parallel-group, placebo-controlled randomized trial in which participants were randomized to one of four conditions: an eGo-style e-cigarette paired with 0, 8, or 36 mg/mL nicotine liquid, with tobacco or menthol flavor (participants chose flavor), or a cigarette-shaped plastic tube with no electronics or aerosol, for use as a cigarette substitute. The blood nicotine boost from 36 mg/mL nicotine liquid with this device (13 ng/mL) was approximately similar to that resulting from smoking a cigarette (around 15 ng/mL).^29^

This trial is registered with ClinicalTrials.gov, NCT02342795. The study followed the Consolidated Standards of Reporting Trials (CONSORT) guideline for randomized clinical trials and the study protocol was approved by the Institutional Review Boards of both sites - Pennsylvania State University (Hershey, PA) and Virginia Commonwealth University, (Richmond, VA), United States.

### Population

Adults aged 21–65 years who smoked at least 10 cigarettes per day (CPD), had an expired-air carbon monoxide (CO) reading of >9 parts per million (ppm) at baseline and were interested in reducing their cigarette consumption were enrolled in the study. Participants with significant medical conditions were excluded in the study to ensure safety, including pregnancy or breastfeeding, unstable cardiovascular disease within the past 12 months, severe immune, respiratory, renal or hepatic disease, uncontrolled mental illness or substance abuse requiring inpatient treatment within the past six months.

During the study, participants were encouraged to reduce their cigarette smoking by 50% for 2 weeks, then by 75% for 2 weeks, and then to maintain 75% reduction and/or continue to try to reduce for the remainder of the trial to 6 months. After screening, participants attended a randomized visit and completed nine post-randomization follow-up visits, including in-person visits at weeks 4, 8, 12 and 24, and phone-call follow-ups at weeks 1, 2, 8, 16, and 20. A detailed description of the study procedure as well as the primary and secondary outcome results, have been published previously.^11,12,30,31^

### Study outcome measures

The main outcome of interest in the current study was the occurrence of AEs reported by participants during the 6-month intervention period (week 1–24). At each scheduled visit, participants were asked standardized AE trigger questions regarding changes in medication, physical or mental health, or any medical care sought since the last visit. All reported AEs were coded using the National Cancer Institute’s (NCI) common terminology criteria for adverse events (CTCAE) v4.0 safety profiler, which assigns Medical Dictionary for Regulatory Activities (MedDRA) preferred terms, severity grades, and event codes. AEs were assessed as follows:

- Frequency was defined as the number of participants experiencing an event that started or intensified during the intervention period.
- Severity was graded as mild, moderate, severe, life-threatening or fatal to describe the intensity of the event.
- The AE was described as serious if the AE met criteria such as death, life-threatening experience, hospitalization, disability/permanent damage or other medically important events.
- Relatedness (causality) was initially assessed by study staff with final determination made by the medical monitor at each site. Relatedness was classified as unrelated, unlikely, possibly, probably, or definitely related. Events were considered possibly related or higher if there was a reasonable possibility (i.e., >50% likelihood) that the AE was attributable to study product use or study procedures, based on clinical judgement and available evidence. If causality was uncertain, AEs were conservatively treated as related for reporting purposes.

In addition to evaluating all reported AEs, analyses focused on study-related AEs to assess safety signals likely attributable to e-cigarette exposure rather than preexisting conditions or unrelated health events. Study-related AEs were defined as those rated as possibly, probably, or definitely related to either the assigned study product or the study procedures (e.g., smoking reduction requirements), as determined by the medical monitor.

### Statistical analysis

A summary of the overall characteristics of all reported AEs and study-related AEs was performed, stratified by randomized groups. The frequency and proportion (%) of participants reporting each type of AE were calculated for each group. Frequently reported AEs were defined as study-related AEs occurring in ≥1% of participants across all study arms and were selected for further analysis.

To estimate between-group differences in AE risk, we calculated risk differences with corresponding 95% confidence intervals (CIs) using nonparametric bootstrapping via the g-computation (gComp) approach with 1000 resamples. We selected risk difference as the effect measure rather than the relative risk or odds ratio, as several AE categories included zero events in one comparison group, which could result in biased or inflated ratio estimates using generalized linear models.

Fisher’s exact test was used to assess statistical differences in AE occurrence across randomized groups. A two-sided p-value of <0.05 was considered statistically significant. Adjusted p-values for multiple comparisons were calculated using the False Discovery Rate (FDR) method.

All the analyses were conducted separately for all reported AEs and for study-related AEs (those deemed possibly, probably, or definitely related to product use). In line with the study’s objective to assess safety signals attributable to e-cigarette use or nicotine e-cigarette use, the analyses focus on contrasts between e-cigarette groups vs. cigarette substitute group, and between nicotine containing e-cigarette groups versus the non-nicotine (placebo) e-cigarette group. Sensitivity analyses were performed including: (1) analyses restricted to AEs reported within the first month of follow-up, (2) comparisons by flavor choice (menthol vs. tobacco), (3) comparisons of each e-cigarette dose group vs. the cigarette substitute, and (4) comparisons of each nicotine e-cigarette group vs. the non-nicotine (placebo) e-cigarette group.

Cumulative incidence of first study-related AE was estimated using Kaplan-Meier methods to account for differential follow-up due to dropout. Participants were censored at the time of withdrawal or at the end of follow-up if no AE occurred. The risk set at each time point included participants who remained under observation and were event-free. All analyses were conducted using R (version 4.3.1).

## RESULTS

The study randomized 520 smokers to one of the four groups (130 participants per group) between July 22, 2015 and Nov 16, 2017. Baseline characteristics of the randomized groups were similar. Detailed description of the participant characteristics has been published previously.^11,12^ The Consort diagram shows that overall, 332 participants (63.8%) continued to attend through to 24 weeks with no significant between-group difference in dropout rates (supplemental figure 1). A summary of study product use across the randomized arms over the intervention period is presented in supplementary table 1. Product use was generally similar across treatment arms. Compared to the cigarette substitute group, participants in the 36 mg/mL group were using their products more during weeks 8-16, and at week 24 (supplementary table 1).

### AE profiles by randomized groups

Table 1 summarizes the frequency, severity, seriousness, and relatedness of AEs (all and study-related) across randomized groups in the trial. Overall, 66.2% of the participants (n=344/520) reported at least one AE. AE reporting differed significantly across randomized groups, with more common reporting in the nicotine e-cigarette groups (72% and 74%) compared to the cigarette substitute and non-nicotine e-cigarette groups (56% and 62%; p=0.007). The total number of AEs reported was 745, with most (90.3%) graded as mild or moderate. There was one fatal event reported in the 0 mg/mL group (death from suicide) that was determined unrelated to the study. Less than 1 in 20 participants (4.3%) experienced serious AEs, with all serious events deemed unrelated or unlikely to be related to study participation.

**Table 1.**
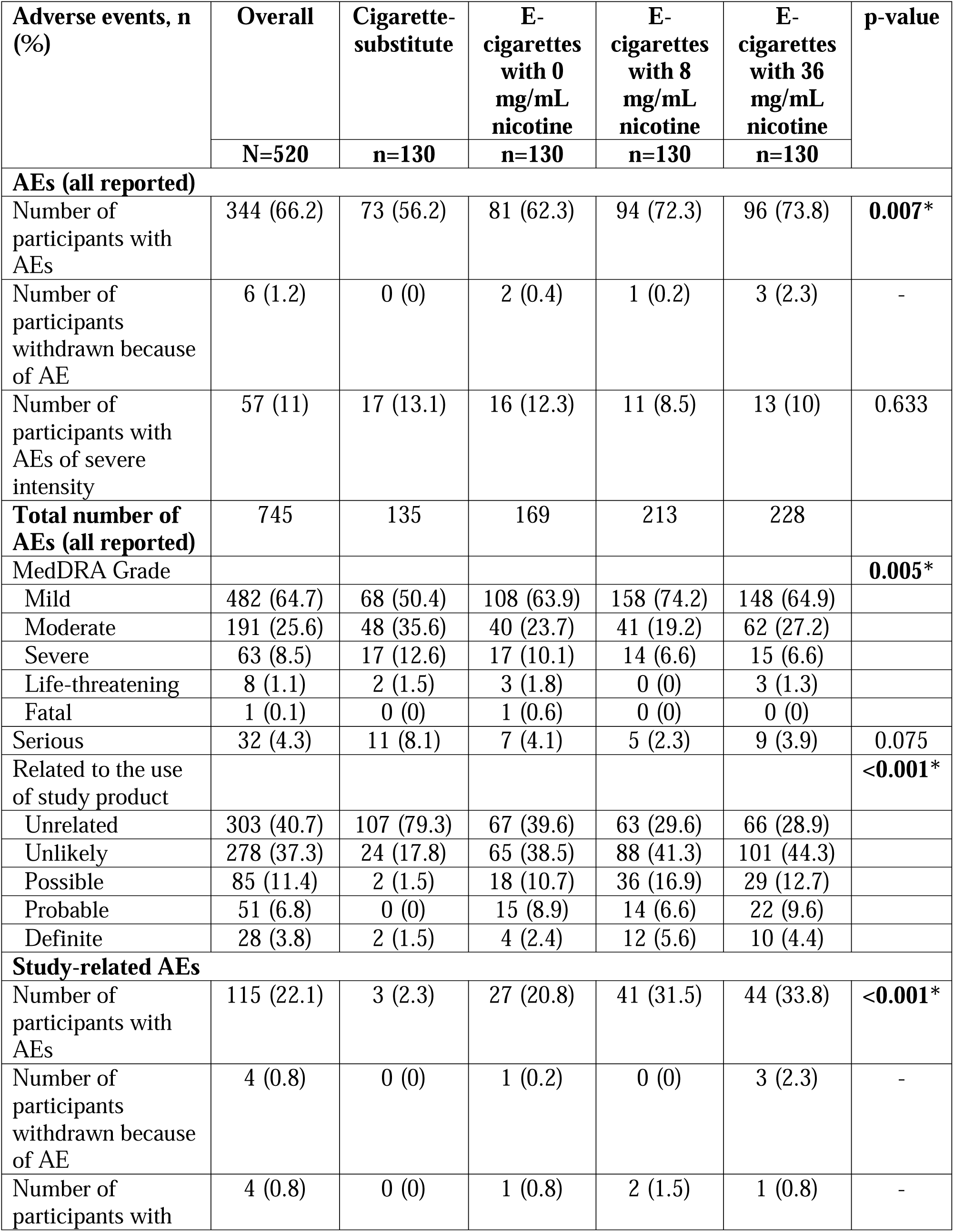
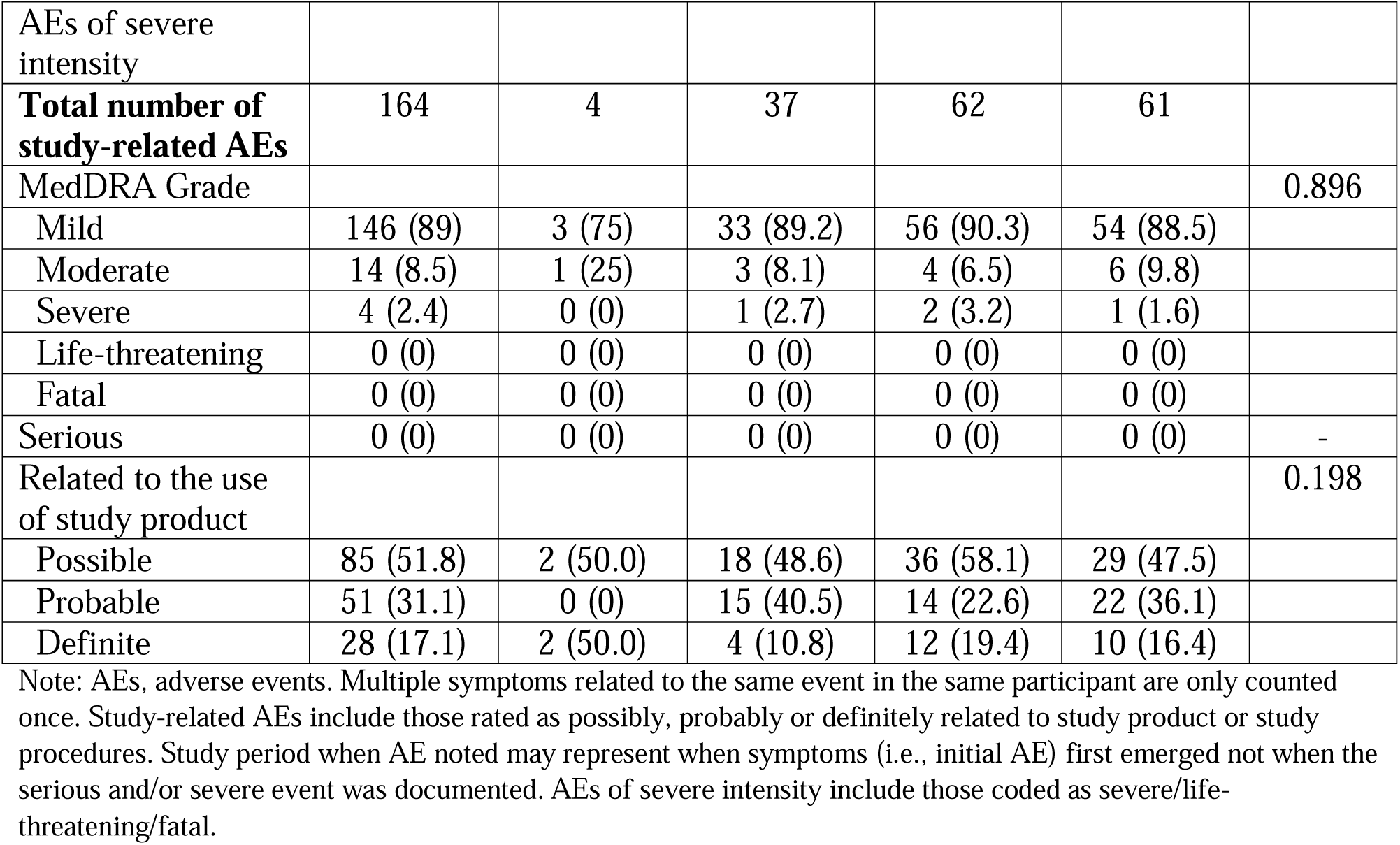
Summary of adverse events overall and by randomized groups.

When restricted to study-related AEs (those rated as possibly, probably, or definitely related to the study product or study procedures), 22.1% of the participants (n=115/520) reported at least one related AE, with significantly higher proportion in the e-cigarette groups (20.8 – 33.8%) compared to cigarette substitute group (2.3%, p<0.001). The total number of study-related AEs reported was 164, with majority of events of mild intensity. Four participants across all groups experienced a severe study-related AE: hypertension (0 mg/mL group), dyspnea (8 mg/mL group), cough and syncope (8 mg/mL group), and cough (36 mg/mL group). There were no serious study-related AEs reported. Also, there were 4 participant withdrawals due to an AE: 3 in 36 mg/mL group (cough, psychiatric – lethargic and spaced out, mouth ulcer), and 1 in 0 mg/mL group (cough and headache) (table 1).

Cumulative incidence of first study-related AE increased over 24 weeks by treatment arms, with higher and earlier accumulation observed in the nicotine e-cigarette groups compared with the 0 mg/mL or cigarette substitute groups (figure 1). By 24 weeks, cumulative incidence was highest in the 36 mg/mL (37.0%) and 8 mg/mL (35.2%) groups, followed by the 0 mg/mL group (23.4%), and lowest in the cigarette substitute group (2.5%). Differences in time to first study-related AE across randomized arms were statistically significant (log-rank p<0.001).

**Figure 1.**
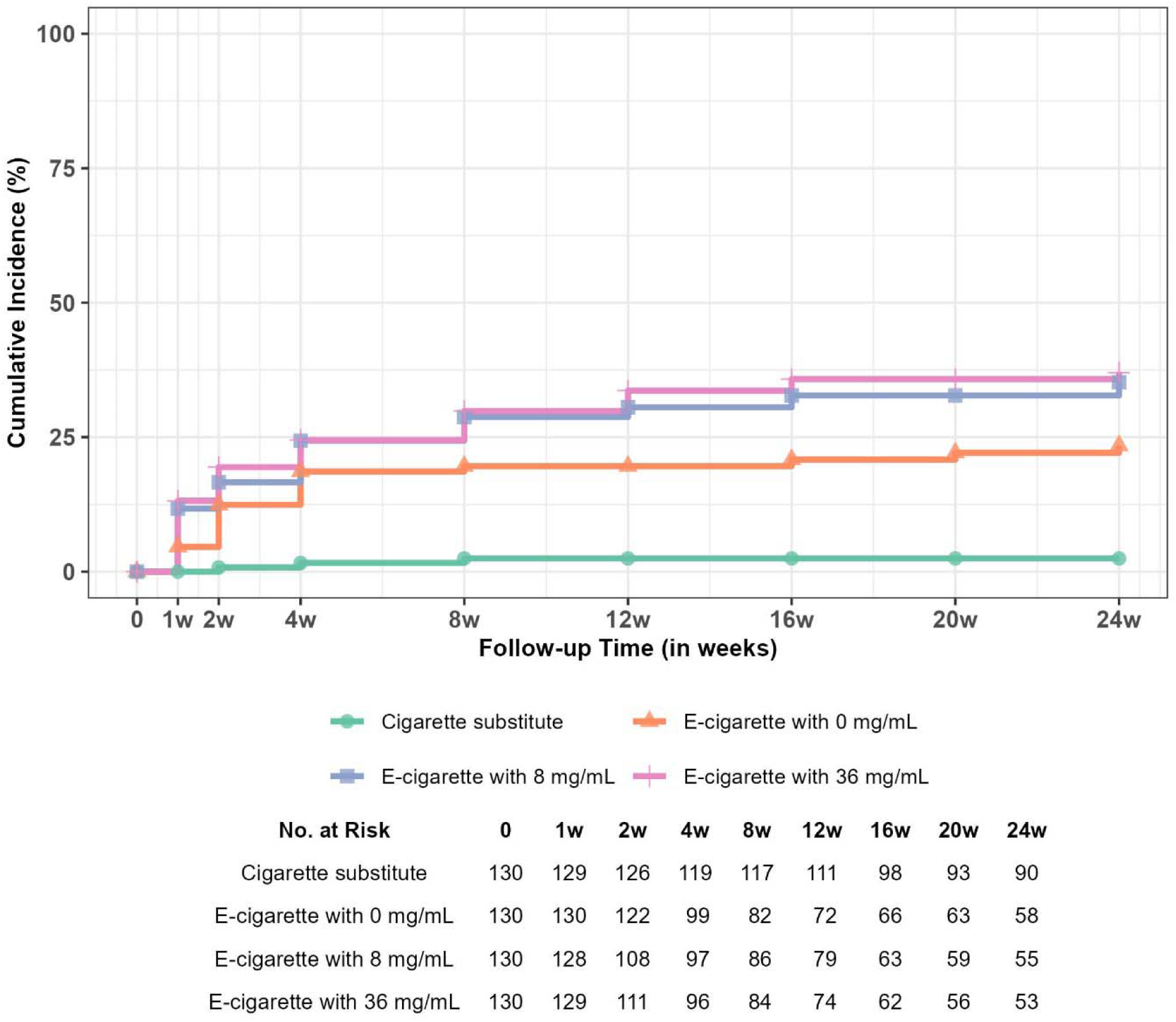
Cumulative incidence of first study-related AEs over the 6-month intervention period by randomized groups. Log-rank test of equality over strata: χ2(3, N = 520) = 46.6, p = <0.001.

### Frequently reported study-related AEs

Detailed description of all and study-related AEs by randomized groups is presented in supplemental tables 2 and 3. The following eight study-related AEs were most frequently reported: cough (6.3%), dizziness (1.0%), dry mouth (1.2%), headache (4.0%), mouth ulcers (2.3%), nausea (2.1%), other respiratory symptoms (e.g., reduction in spirometry indices, chest tightness, or gagging sensation) (1.2%), and sore throat (4.0%) (supplemental table 2). Except for dry mouth, all above-mentioned study-related AEs were frequently reported in the first month (supplemental table 4 and 5). For each frequently reported study-related AEs, most first occurrences were reported as mild, and AEs of severe intensity were uncommon (Table 2).

**Table 2.** Counts of participants in each level of severity for first occurrence of most frequently reported study-related AEs^a,b,c^.

| Adverse events,<br>n (%) | Overall | Cigarette-substitute | E-cigarettes<br>with 0<br>mg/mL<br>nicotine | E-cigarettes<br>with 8<br>mg/mL<br>nicotine | E-cigarettes<br>with 36<br>mg/mL<br>nicotine |
| --- | --- | --- | --- | --- | --- |
|  | N=520 | n=130 | n=130 | n=130 | n=130 |
| **Cough** | 33 (6.3) | 0 (0) | 4 (3.1) | 18 (13.8) | 11 (8.5) |
| Mild | 31 (6.0) | 0 (0) | 4 (3.1) | 17 (13.1) | 10 (7.7) |
| Moderate | 0 (0) | 0 (0) | 0 (0) | 0 (0) | 0 (0) |
| Severe | 2 (0.4) | 0 (0) | 0 (0) | 1 (0.8) | 1 (0.8) |
| **Dizziness** | 5 (1.0) | 0 (0) | 2 (1.5) | 2 (1.5) | 1 (0.8) |
| Mild | 4 (0.8) | 0 (0) | 2 (1.5) | 1 (0.8) | 1 (0.8) |
| Moderate | 1 (0.2) | 0 (0) | 0 (0) | 1 (0.8) | 0 (0) |
| Severe | 0 (0) | 0 (0) | 0 (0) | 0 (0) | 0 (0) |
| **Dry mouth** | 6 (1.2) | 0 (0) | 1 (0.8) | 2 (1.5) | 3 (2.3) |
| Mild | 6 (1.2) | 0 (0) | 1 (0.8) | 2 (1.5) | 3 (2.3) |
| Moderate | 0 (0) | 0 (0) | 0 (0) | 0 (0) | 0 (0) |
| Severe | 0 (0) | 0 (0) | 0 (0) | 0 (0) | 0 (0) |
| **Headache** | 21 (4.0) | 0 (0) | 7 (5.4) | 7 (5.4) | 7 (5.4) |
| Mild | 20 (3.8) | 0 (0) | 7 (5.4) | 7 (5.4) | 6 (4.6) |
| Moderate | 1 (0.2) | 0 (0) | 0 (0) | 0 (0) | 1 (0.8) |
| Severe | 0 (0) | 0 (0) | 0 (0) | 0 (0) | 0 (0) |
| **Mouth ulcers** | 12 (2.3) | 0 (0) | 2 (1.5) | 5 (3.8) | 5 (3.8) |
| Mild | 12 (2.3) | 0 (0) | 2 (1.5) | 5 (3.8) | 5 (3.8) |
| Moderate | 0 (0) | 0 (0) | 0 (0) | 0 (0) | 0 (0) |
| Severe | 0 (0) | 0 (0) | 0 (0) | 0 (0) | 0 (0) |
| **Nausea** | 11 (2.1) | 0 (0) | 2 (1.5) | 4 (3.1) | 5 (3.8) |
| Mild | 10 (2.1) | 0 (0) | 1 (0.8) | 4 (3.1) | 5 (3.8) |
| Moderate | 1 (0.2) | 0 (0) | 1 (0.8) | 0 (0) | 0 (0) |
| Severe | 0 (0) | 0 (0) | 0 (0) | 0 (0) | 0 (0) |
| **Other respiratory**<br>**symptoms** | 6 (1.2) | 0 (0) | 0 (0) | 0 (0) | 6 (4.6) |
| Mild | 6 (1.2) | 0 (0) | 0 (0) | 0 (0) | 6 (4.6) |
| Moderate | 0 (0) | 0 (0) | 0 (0) | 0 (0) | 0 (0) |
| Severe | 0 (0) | 0 (0) | 0 (0) | 0 (0) | 0 (0) |
| **Sore throat** | 21 (4.0) | 0 (0) | 6 (4.6) | 5 (3.8) | 10 (7.7) |
| Mild | 21 (4.0) | 0 (0) | 6 (4.6) | 5 (3.8) | 10 (7.7) |
| Moderate | 0 (0) | 0 (0) | 0 (0) | 0 (0) | 0 (0) |
| Severe | 0 (0) | 0 (0) | 0 (0) | 0 (0) | 0 (0) |

<sup>a</sup> AE, adverse events; Counts represent the number of participants reporting the first occurrence of each AE at the specified severity level.
<sup>b</sup> Severity was classified according to MedDRA intensity grades (mild, moderate, severe, life-threatening, fatal).
<sup>c</sup> Participants are counted once per AE using the maximum intensity at first occurrence.

**Table 3.** Estimated risk differences for frequently reported study-related adverse events: E-cigarette vs. cigarette substitute groups.

| Adverse events | Cigarette substitute | E-cigarettes with 0/8/36 mg/mL nicotine | p-value<sup>†</sup> | Risk difference (CI), % |
| --- | --- | --- | --- | --- |
| **Yes, n (%)** | **n = 120** | **n = 390** |  |  |
| Cough | 0 (0) | 33 (8.5) | **<0.001\*** | 8.5 (5.6 – 11.3) |
| Dizziness | 0 (0) | 5 (1.3) | 0.345 | 1.3 (0.3 – 2.5) |
| Dry mouth | 0 (0) | 6 (1.5) | 0.345 | 1.5 (0.5 – 2.9) |
| Headache | 0 (0) | 21 (5.4) | **0.008\*** | 5.4 (3.3 – 7.6) |
| Mouth ulcers | 0 (0) | 12 (3.1) | 0.088 | 3.1 (1.3 – 4.9) |
| Nausea | 0 (0) | 11 (2.8) | 0.118 | 2.8 (1.3 – 4.6) |
| Other respiratory symptoms | 0 (0) | 6 (1.5) | 0.345 | 1.5 (0.3 – 3.0) |
| Sore throat | 0 (0) | 21 (5.4) | **0.008\*** | 5.4 (3.2 – 7.6) |

Note: Study-related adverse events include those rated as possibly, probably or definitely related to study product or study procedures.
Reference group: Cigarette substitute.
p-values reported from Fisher exact test.
\* $p < 0.05$ ; <sup>†</sup>FDR-adjusted p-values.

### Risk differences in frequently reported AEs: E-cigarette vs cigarette substitute groups

Estimated risk differences for frequently reported study-related AEs between e-cigarette and cigarette substitute groups are presented in table 3 and supplemental table 6. Participants randomized to e-cigarette groups reported a significantly higher incidence of cough compared to those in the cigarette substitute group (8.5% vs 0%; p<0.001). The risk of experiencing headache and sore throat was also significantly higher in the e-cigarette group, with both events showing a risk difference of 5.4 percentage points relative to the cigarette substitute group (p=0.008 for both; table 3).

As shown in supplemental tables 7 and 8, similar patterns were observed when evaluating all AEs, regardless of relatedness, between e-cigarette groups and the cigarette substitute group (i.e., e-cigarette groups had significantly higher rates of AEs relative to the cigarette substitute group). Additionally, sensitivity analyses restricted to AEs reported (all and study-related) within the first month of follow-up showed comparable results (supplemental tables 9 – 12).

### Risk differences in frequently reported AEs: Nicotine vs non-nicotine e-cigarette groups

Participants randomized to nicotine e-cigarette groups experienced a significantly higher incidence of cough compared to the non-nicotine e-cigarette group (11.2% vs 3.1%; p=0.048; table 4). When analyses were stratified by nicotine concentration, the incidence of cough was significantly higher in the 8 mg/mL group only compared to the 0 mg/mL group (13.8% vs 3.1%; p=0.003), suggesting no clear dose-response relationship (supplemental table 13).

**Table 4.** Estimated risk differences for frequently reported study-related adverse events: e-cigarette with 0 vs. 8/36 mg/mL nicotine groups.

| Adverse events | E-cigarette with 0 mg/mL nicotine | E-cigarettes with 8/36 mg/mL nicotine | p-value<sup>†</sup> | Risk difference (CI), % |
| --- | --- | --- | --- | --- |
| **Yes, n (%)** | **n= 120** | **n = 240** |  |  |
| Cough | 4 (3.1) | 29 (11.2) | **0.048\*** | 8.1<br>(3.4 – 12.8) |
| Dizziness | 2 (1.5) | 3 (1.2) | 1 | -0.4<br>(-3.1 – 2.0) |
| Dry mouth | 1 (0.8) | 5 (1.9) | 1 | 1.1<br>(-0.9 – 3.4) |
| Headache | 7 (5.4) | 14 (5.4) | 1 | 0<br>(-5 – 4.5) |
| Mouth ulcers | 2 (1.5) | 10 (3.8) | 0.702 | 2.3<br>(-1.1 – 5.2) |
| Nausea | 2 (1.5) | 9 (3.5) | 0.702 | 1.9<br>(-1.2 – 5.0) |
| Other respiratory symptoms | 0 (0) | 6 (2.3) | 0.702 | 2.3<br>(0.8 – 4.4) |
| Sore throat | 6 (4.6) | 15 (5.8) | 1 | 1.1<br>(-3.8 – 5.3) |

Note: Study-related adverse events include those rated as possibly, probably or definitely related to study product or study procedures.
Reference group: E-cigarette with 0 mg/mL nicotine.
p-values reported from Fisher exact test.
\* $p<0.05$ ; <sup>†</sup>FDR-adjusted p-values.

Additionally, six participants in the 36 mg/mL group had other respiratory symptoms such as reductions in spirometry indices, chest tightness, or gagging sensation, which was significantly higher compared to the 0 mg/mL group (4.6% vs. 0%; p=0.030; supplemental table 13). No other frequently reported study-related AEs differed significantly between nicotine and non-nicotine e-cigarette groups (table 4 and supplemental table 13).

When evaluating all AEs regardless of relatedness, cough remained significantly more common among nicotine e-cigarette groups compared to the non-nicotine e-cigarette groups (supplemental tables 14 and 15). Analyses restricted to AEs reported during the first month of follow-up showed similar patterns for both all and study-related AEs (supplemental tables 16 – 19).

Finally, we also explored AE reporting by flavor choice and found no significant differences in AE incidence between participants using menthol- versus tobacco-flavored e-cigarettes (supplemental table 20). Stratified analyses by nicotine concentration in e-cigarette arms likewise did not reveal consistent differences in AE incidence by flavor choice (data not shown).

## DISCUSSION

Safety evaluations from RCTs of e-cigarettes have been limited, with most prior studies focusing primarily on serious adverse events (SAEs) as secondary outcomes.^11,16,32^ To date, this study is among the first to directly compare AE profiles across e-cigarettes with varying nicotine concentrations and a cigarette substitute in a double-blind, 6-month trial designed to reflect real-world use among individuals wanting to reduce combustible cigarette use. Across groups, the study found no serious study-related AEs and few study withdrawals due to AEs (4/520 participants), suggesting that all study products were generally well tolerated during the intervention period. This finding is consistent with several RCTs and systematic reviews, which report that most AEs associated with e-cigarette use are non-serious.^10,33–36^

Participants assigned to e-cigarette groups reported a significantly higher incidence of study-related AEs compared to the cigarette substitute group. These differences are likely attributable to the inhalation of aerosolized constituents such as PG and VG, which can produce toxicants, including carbonyl compounds and reactive free radicals,^37^ associated with inflammatory and oxidative stress responses, and DNA damage.^38^ The incidence of AEs was even significantly higher among those assigned to nicotine-containing e-cigarettes, suggesting that nicotine may exert additive effects on symptom burden consistent with its known potential to adversely affect respiratory and cardiovascular systems.^39^

The nicotine-containing e-cigarette groups showed a significantly higher incidence of cough compared to the non-nicotine group. While we did not observe a clear dose-response relationship across the 8 mg/mL and 36 mg/mL arms, findings align with conclusions drawn by the National Academies of Sciences, Engineering, and Medicine (NASEM), which highlighted that nicotine-containing e-cigarettes but not non-nicotine e-cigarettes could have short-term adverse effects on lung defense mechanisms such as mucociliary clearance, urge to cough and cough sensitivity.^13^ In addition, users assigned to higher nicotine concentrations of e-cigarettes may shorten puff duration^29^ in response to the sensory characteristics of the aerosol, potentially attenuating an expected dose-response relationship. Furthermore, the increase in other respiratory symptoms (e.g., reductions in spirometry indices, chest tightness, or gagging sensation) seen only among participants randomized to the 36 mg/mL group supports that higher inhaled nicotine exposure may be linked to acute respiratory effects. However, an RCT involving cigarette users (>10/day) which also used pen-style e-cigarettes (16 mg and 0 mg) showed mouth/throat irritation and nausea as the most commonly reported adverse event, with no reports of cough.^40^ These findings highlight the variability in AE reporting based on device characteristics and liquid composition, emphasizing the importance of continued safety evaluations of e-cigarette aerosol in both clinical and preclinical settings.

Critically, the findings from this study should be considered in light of the study context. This study was primarily a smoking reduction trial, and the majority of the participants continued to smoke while using their assigned study products, resulting in a dual-use pattern. Dual use of combustible cigarettes and e-cigarettes has been a subject of ongoing public health concern, particularly due to the potential for nicotine toxicity and increased toxicant exposure.^41^ In this context, adverse events that might be anticipated include symptoms consistent with nicotine excess, such as headache, nausea, dizziness, and vomiting among participants assigned to nicotine e-cigarette groups.^42^ However, our findings indicate that such AEs were infrequent across all groups, including those assigned to 36 mg/mL nicotine e-cigarettes, which were capable of delivering nicotine levels approaching those of combustible cigarettes. The only frequently reported AEs with plausible links to nicotine exposure were headache and nausea, both of which were generally mild in severity. These findings may support the short-term safety and tolerability of nicotine e-cigarette use, particularly in the context of harm reduction for individuals who smoke.

The most frequently reported study-related AEs in this trial were – cough, dizziness, dry mouth, headache, mouth ulcers, nausea, other respiratory symptoms, and sore throat, most of which align with those identified in prior research.^43–47^ Interestingly, nicotine withdrawal symptoms such as anxiety, irritability, and insomnia were not commonly reported, possibly due to the nicotine replacement provided by the e-cigarettes and concurrent use of combustible cigarettes while reducing during the intervention period. Findings also demonstrated a higher risk of cough, headache, and sore throat in e-cigarette groups compared to those in cigarette substitute group.

These results are consistent with recent systematic reviews and meta-analyses indicating that mild AEs are more common following e-cigarette use relative to behavioral support only or no support,^10^ but these rates are not significantly different when compared to NRT^10,19^ or non-nicotine e-cigarettes.^10,17,19^ Findings from this trial may help clinicians counsel and communicate the AE profile of e-cigarette use to patients who have been unsuccessful in smoking cessation using FDA-approved therapies and are considering e-cigarettes as an alternative.

The major strengths of this study include placebo-controlled design including an additional randomized control group who did not use an e-cigarette device, a large and a diverse sample, and a relatively long follow-up period (6 months). However, the trial also had several limitations. First, the cigarette substitute arm could not be blinded. This design detail could partially explain the low number of study-related AEs observed for this group and may have inflated the observed risk differences when compared to the e-cigarette groups. Nevertheless, several sensitivity analyses such as those including all reported AEs regardless of relatedness, and analysis restricted to the first month of follow-up showed patterns consistent with primary findings.

Additionally, the study utilized second-generation free-base nicotine e-cigarettes, which differ from newer nicotine salt-based formulations in terms of delivery profile and user experience. However, the primary constituents such as PG/VG, nicotine, and flavorings are common to most e-cigarettes in current use, suggesting that the safety signals identified in this study remain relevant across products. Finally, participants were screened for eligibility and required to meet specific health and safety criteria prior to enrollment. As such, the safety profile observed in this trial may not fully reflect real-world use, where individuals with unstable or significant health conditions (e.g., exacerbations of asthma or COPD, or uncontrolled hypertension) may experience different or higher rates of AEs.

## CONCLUSION

This RCT provides one of the most comprehensive evaluations of AE profiles across e-cigarette products with varying nicotine concentrations and a non-nicotine, non-aerosol cigarette substitute. While study-related AEs such as cough, headache, and sore throat were more commonly reported among participants using e-cigarettes, especially those containing nicotine, the majority were mild in nature and did not lead to serious health concerns. These findings support the short-term safety and tolerability of e-cigarette use for individuals wanting to reduce combustible cigarette use. Future research should include long-term studies with standardized AE reporting across evolving product types and diverse user populations to better inform clinical and regulatory decision-making.

## DISCLAIMER

The content is solely the responsibility of the authors and does not necessarily represent the official views of the National Institutes of Health, or the U.S. Food and Drug Administration.

## DATA AVAILABILITY STATEMENT

Data are available on reasonable request. On publication, requests for deidentified individual participant data or study documents (e.g., data dictionary; protocol; statistical analysis plan; measures, manuals or informed consent documentation) will be considered. The requestor must submit a one-page abstract of their proposed research, including the purpose, analytical plan and dissemination plans. The Executive Leadership Committee (Virginia Commonwealth University and Penn State University) will review the abstract and decide on the basis of the individual merits. Review criteria and prioritization of projects include potential of the proposed work to advance public health, qualifications of the applicant, the potential for publication, the potential for future funding and enhancing the scientific, geographic and demographic diversity of the research portfolio. Following abstract approval, requestors must receive institutional ethics approval or confirmation of exempt status for the proposed research. An executed data use agreement must be completed before data distribution. Requests should be made to the corresponding author.

## ACKNOWLEDGEMENTS

The project described was supported by the National Center for Advancing Translational Sciences, National Institutes of Health, through Grant UL1TR002014, which supports the Penn State Clinical and Translational Science Institute and its Clinical Research Centre, where participant visits were conducted and Grant UL1TR002649 at Virginia Commonwealth University where data was collected.

## REFERENCES

1. U.S. Department of Health and Human Services. The Health Consequences of Smoking: 50 Years of Progress. A Report of the Surgeon General. Atlanta, GA: U.S. Department of Health and Human Services, Centers for Disease Control and Prevention, National Center for Chronic Disease Prevention and Health Promotion, Office on Smoking and Health, 2014.

2. CDC. Cigarette Smoking. Smoking and Tobacco Use. January 6, 2025. Accessed December 14, 2025. https://www.cdc.gov/tobacco/about/index.html

3. Benowitz NL. Nicotine Addiction. New England Journal of Medicine. 2010;362(24):2295–2303. doi:10.1056/NEJMra0809890

4. Panel TU and DG. Treating Tobacco Use and Dependence: 2008 Update. US Department of Health and Human Services; 2008.

5. Cahill K, Stevens S, Perera R, Lancaster T. Pharmacological interventions for smoking cessation: an overview and network meta analysis - Cahill, K - 2013 | Cochrane Library. Accessed December 14, 2025. https://www.cochranelibrary.com/cdsr/doi/10.1002/14651858.CD009329.pub2/full

6. Shiffman S, Brockwell SE, Pillitteri JL, Gitchell JG. Individual differences in adoption of treatment for smoking cessation: Demographic and smoking history characteristics. Drug and Alcohol Dependence. 2008;93(1):121–131. doi:10.1016/j.drugalcdep.2007.09.005

7. Levy DT, Cummings KM, Villanti AC, et al. A framework for evaluating the public health impact of e-cigarettes and other vaporized nicotine products. Addiction. 2017;112(1):8–17. doi:10.1111/add.13394

8. Breland A, Soule E, Lopez A, Ramôa C, El-Hellani A, Eissenberg T. Electronic cigarettes: what are they and what do they do? Annals of the New York Academy of Sciences. 2017;1394(1):5–30. doi:10.1111/nyas.12977

9. McNeill A, Brose L, Robson D, et al. Nicotine vaping in England: An evidence update including health risks and perceptions, 2022. Published online September 29, 2022. Accessed March 23, 2023. https://kclpure.kcl.ac.uk/portal/en/publications/nicotine-vaping-in-england(58869843-3d0c-40e7-a9e9-d92881a2ed22).html

10. Lindsona N, Livingstone-Banksa J, Butler AR, et al. Electronic cigarettes for smoking cessation. Cochrane Database of Systematic Reviews. 2025;2025(11). doi:10.1002/14651858.cd010216.pub10

11. Cobb CO, Foulds J, Yen MS, et al. Effect of an electronic nicotine delivery system with 0, 8, or 36 mg/mL liquid nicotine versus a cigarette substitute on tobacco-related toxicant exposure: a four-arm, parallel-group, randomised, controlled trial. The Lancet Respiratory Medicine. 2021;9(8):840–850. doi:10.1016/S2213-2600(21)00022-9

12. Foulds J, Cobb CO, Yen MS, et al. Effect of Electronic Nicotine Delivery Systems on Cigarette Abstinence in Smokers With No Plans to Quit: Exploratory Analysis of a Randomized Placebo-Controlled Trial. Nicotine Tob Res. 2022;24(7):955–961. doi:10.1093/ntr/ntab247

13. National Academies of Sciences, Engineering, and Medicine. Public Health Consequences of E-Cigarettes. Washington, DC, The National Academies Press, 2018.

14. Rose JJ, Krishnan-Sarin S, Exil VJ, et al. Cardiopulmonary Impact of Electronic Cigarettes and Vaping Products: A Scientific Statement From the American Heart Association. Circulation. 2023;148(8):703–728. doi:10.1161/CIR.0000000000001160

15. Thomas KH, Dalili MN, López-López JA, et al. Comparative clinical effectiveness and safety of tobacco cessation pharmacotherapies and electronic cigarettes: a systematic review and network meta-analysis of randomized controlled trials. Addiction. 2022;117(4):861–876. doi:10.1111/add.15675

16. Levett JY, Filion KB, Reynier P, Prell C, Eisenberg MJ. Efficacy and Safety of E-Cigarette Use for Smoking Cessation: A Systematic Review and Meta-Analysis of Randomized Controlled Trials. The American Journal of Medicine. 2023;136(8):804–813.e4. doi:10.1016/j.amjmed.2023.04.014

17. Vyas N, Bennett A, Hamel C, et al. Effectiveness of e-cigarettes as a stop smoking intervention in adults: a systematic review. Syst Rev. 2024;13(1):168. doi:10.1186/s13643-024-02572-7

18. Manzoli L, Flacco ME, Ferrante M, et al. Cohort study of electronic cigarette use: effectiveness and safety at 24 months. Tobacco Control. 2017;26(3):284–292. doi:10.1136/tobaccocontrol-2015-052822

19. Anandan AS, Leung J, Chan GCK, et al. Common adverse events of electronic cigarettes compared with traditional nicotine replacement therapies: A systematic review and meta-analysis. Drug and Alcohol Review. 2023;42(5):1278–1287. doi:10.1111/dar.13674

20. FDA. Premarket Tobacco Product Applications. March 27, 2024. Accessed December 15, 2025. https://www.fda.gov/tobacco-products/market-and-distribute-tobacco-product/premarket-tobacco-product-applications

21. Motooka Y, Matsui T, Slaton RM, et al. Adverse events of smoking cessation treatments (nicotine replacement therapy and non-nicotine prescription medication) and electronic cigarettes in the Food and Drug Administration Adverse Event Reporting System, 2004−2016. Sage Open Medicine. 2018;6:2050312118777953. doi:10.1177/2050312118777953

22. Lindson N, Theodoulou A, Ordóñez-Mena JM, et al. Pharmacological and electronic cigarette interventions for smoking cessation in adults: component network meta analyses - Lindson, N - 2023 | Cochrane Library. Accessed December 15, 2025. https://www.cochranelibrary.com/cdsr/doi/10.1002/14651858.CD015226.pub2/full

23. de Dios MA, Anderson BJ, Stanton C, Audet DA, Stein M. Project Impact: a pharmacotherapy pilot trial investigating the abstinence and treatment adherence of Latino light smokers. J Subst Abuse Treat. 2012;43(3):322–330. doi:10.1016/j.jsat.2012.01.004

24. Balmford J, Borland R, Hammond D, Cummings KM. Adherence to and Reasons for Premature Discontinuation From Stop-Smoking Medications: Data From the ITC Four-Country Survey. Nicotine Tob Res. 2011;13(2):94–102. doi:10.1093/ntr/ntq215

25. Raupach T, Brown J, Herbec A, Brose L, West R. A systematic review of studies assessing the association between adherence to smoking cessation medication and treatment success. Addiction. 2014;109(1):35–43. doi:10.1111/add.12319

26. FDA. E-Cigarettes, “Vapes” and Other Electronic Nicotine Delivery Systems (ENDS) Authorized by the FDA. Published online September 18, 2025. Accessed December 15, 2025. https://www.fda.gov/tobacco-products/market-and-distribute-tobacco-product/e-cigarettes-vapes-and-other-electronic-nicotine-delivery-systems-ends-authorized-fda

27. Gotts JE, Jordt SE, McConnell R, Tarran R. What are the respiratory effects of e-cigarettes? BMJ. 2019;366:l5275. doi:10.1136/bmj.l5275

28. Kabele M, Lyytinen G, Bosson JA, et al. Nicotine in E-Cigarette Aerosol May Lead to Pulmonary Inflammation. SSRN. Preprint posted online 2025. doi:10.2139/ssrn.5122926

29. Hiler M, Breland A, Spindle T, et al. Electronic Cigarette User Plasma Nicotine Concentration, Puff Topography, Heart Rate, and Subjective Effects: Influence of Liquid Nicotine Concentration and User Experience. Exp Clin Psychopharmacol. 2017;25(5):380–392. doi:10.1037/pha0000140

30. Lopez AA, Cobb CO, Yingst JM, et al. A transdisciplinary model to inform randomized clinical trial methods for electronic cigarette evaluation. BMC Public Health. 2016;16(1):217. doi:10.1186/s12889-016-2792-8

31. Dahal S, Yingst J, Wang X, et al. Changes in cardiovascular disease risk, lung function and other clinical health outcomes when people who smoke use e-cigarettes to reduce cigarette smoking: an exploratory analysis from a randomised placebo-controlled trial. BMJ Open. 2025;15(6):e098005. doi:10.1136/bmjopen-2024-098005

32. Bonevski B, Rich J, Lubman DI, et al. Nicotine e-cigarettes for smoking cessation following discharge from smoke-free inpatient alcohol and other drug withdrawal services: a pragmatic two-arm, single-blinded, parallel-group, randomised controlled trial. The Lancet Public Health. 2025;10(7):e568–e577. doi:10.1016/S2468-2667(25)00101-X

33. Caponnetto P, Campagna D, Cibella F, et al. EffiCiency and Safety of an eLectronic cigAreTte (ECLAT) as Tobacco Cigarettes Substitute: A Prospective 12-Month Randomized Control Design Study. PLOS ONE. 2013;8(6):e66317. doi:10.1371/journal.pone.0066317

34. Hajek P, Phillips-Waller A, Przulj D, et al. A Randomized Trial of E-Cigarettes versus Nicotine-Replacement Therapy. New England Journal of Medicine. 2019;380(7):629–637. doi:10.1056/NEJMoa1808779

35. Pulvers K, Nollen NL, Rice M, et al. Effect of Pod e-Cigarettes vs Cigarettes on Carcinogen Exposure Among African American and Latinx Smokers: A Randomized Clinical Trial. JAMA Netw Open. 2020;3(11):e2026324. doi:10.1001/jamanetworkopen.2020.26324

36. D’Ruiz CD, Graff DW, Robinson E. Reductions in biomarkers of exposure, impacts on smoking urge and assessment of product use and tolerability in adult smokers following partial or complete substitution of cigarettes with electronic cigarettes. BMC Public Health. 2016;16(1):543. doi:10.1186/s12889-016-3236-1

37. Bitzer ZT, Goel R, Reilly SM, et al. Effects of Solvent and Temperature on Free Radical Formation in Electronic Cigarette Aerosols. Chem Res Toxicol. 2018;31(1):4–12. doi:10.1021/acs.chemrestox.7b00116

38. Sun Y, Lin K, Effah F, et al. Toxicity of humectants propylene glycol and vegetable glycerin in electronic nicotine delivery systems. Toxicol Lett. 2025;413:111739. doi:10.1016/j.toxlet.2025.111739

39. La Rosa G, Vernooij R, Qureshi M, Polosa R, O’Leary R. Clinical testing of the cardiovascular effects of e-cigarette substitution for smoking: a living systematic review. Intern Emerg Med. 2023;18(3):917–928. doi:10.1007/s11739-022-03161-z

40. Bullen C, McRobbie H, Thornley S, Glover M, Lin R, Laugesen M. Effect of an electronic nicotine delivery device (e cigarette) on desire to smoke and withdrawal, user preferences and nicotine delivery: randomised cross-over trial. Tobacco Control. 2010;19(2):98–103. doi:10.1136/tc.2009.031567

41. Goniewicz ML, Smith DM, Edwards KC, et al. Comparison of Nicotine and Toxicant Exposure in Users of Electronic Cigarettes and Combustible Cigarettes. JAMA Netw Open. 2018;1(8):e185937. doi:10.1001/jamanetworkopen.2018.5937

42. Henstra C, Dekkers BGJ, Olgers TJ, ter Maaten JC, Touw DJ. Managing intoxications with nicotine-containing e-liquids. Expert Opinion on Drug Metabolism & Toxicology. 2022;18(2):115–121. doi:10.1080/17425255.2022.2058930

43. Myers Smith K, Phillips-Waller A, Pesola F, et al. E-cigarettes versus nicotine replacement treatment as harm reduction interventions for smokers who find quitting difficult: randomized controlled trial. Addiction. 2022;117(1):224–233. doi:10.1111/add.15628

44. Lee SH, Ahn SH, Cheong YS. Effect of Electronic Cigarettes on Smoking Reduction and Cessation in Korean Male Smokers: A Randomized Controlled Study. J Am Board Fam Med. 2019;32(4):567–574. doi:10.3122/jabfm.2019.04.180384

45. La Rosa GRM, Del Giovane C, Minozzi S, et al. Oral health effects of non combustible nicotine products: a systematic review and network meta analysis of randomized controlled trials. Journal of Dentistry. 2025;160:105910. doi:10.1016/j.jdent.2025.105910

46. Zakiyah N, Purwadi FV, Insani WN, et al. Effectiveness and Safety Profile of Alternative Tobacco and Nicotine Products for Smoking Reduction and Cessation: A Systematic Review. JMDH. 2021;14:1955–1975. doi:10.2147/JMDH.S319727

47. Güttinger E, Mosimann A, Tal K, et al. Adverse symptoms attributed to e-cigarettes over six months among participants of a randomized controlled trial testing nicotine freebase e-cigarettes for smoking cessation – Secondary analysis of the ESTxENDS trial. Nicotine Tob Res. Published online February 20, 2026:ntag038. doi:10.1093/ntr/ntag038

