## Supplemental figure 1, Supplemental table 1 - 20. for "Safety and tolerability of electronic cigarettes to reduce cigarette smoking: Secondary analysis from a randomized placebo-controlled trial"

### Supplemental Files

Supplemental figure 1: Consort Flow Diagram	3

Supplemental table 1. Study product use percentages using intent-to-treat method (with assumption of no use to missing study product usage logs)	5

Supplemental table 2. Study-related adverse events by randomized groups	6

Supplemental table 3. Adverse Events by randomized groups	8

Supplemental table 4. Study-related adverse events reported in the first month by randomized groups	12

Supplemental table 5. Adverse Events reported in the first month by randomized groups	14

Supplemental table 6. Estimation of risk difference of frequently reported study-related adverse events by randomized groups	17

Supplemental table 7. Estimation of risk difference of frequently reported adverse events: e-cigarette vs. cigarette substitute groups	19

Supplemental table 8. Estimation of risk difference of frequently reported adverse events by randomized groups	20

Supplemental table 9. Estimation of risk difference of frequently reported study-related adverse events in the first month: e-cigarette vs. cigarette substitute groups	22

Supplemental table 10. Estimation of risk difference of frequently reported study-related adverse events in the first month by randomized groups	23

Supplemental table 11. Estimation of risk difference of frequently reported adverse events in the first month: e-cigarette vs. cigarette substitute groups	25

Supplemental table 12. Estimation of risk difference of frequently reported adverse events in the first month by randomized groups	26

Supplemental table 13. Estimation of risk difference of frequently reported study-related adverse events by e-cigarette groups	28

Supplemental table 14. Estimation of risk difference of frequently reported adverse events: e-cigarette with 0 vs. 8/36 mg/mL nicotine groups	30

Supplemental table 15. Estimation of risk difference of frequently reported adverse events by e-cigarette groups	31

Supplemental table 16. Estimation of risk difference of frequently reported study-related adverse events in the first month: e-cigarette with 0 vs. 8/36 mg/mL nicotine groups	33

Supplemental table 17. Estimation of risk difference of frequently reported study-related adverse events in the first month by e-cigarette groups	34

Supplemental table 18. Estimation of risk difference of frequently reported adverse events in the first month: e-cigarette with 0 vs. 8/36 mg/mL nicotine groups	36

Supplemental table 19. Estimation of risk difference of frequently reported adverse events in the first month by e-cigarette arms	37

Supplemental table 20. Study-related adverse events by flavor choice among participants randomized to e-cigarette arms	39

Supplemental figure 1: Consort Flow Diagram

2009 participants assessed for eligibility

684 screened in-person at w1

1325 excluded

716 ineligible

98 eligible did not schedule screening

511 no show for screening

520 randomized at w0

130 assigned
cigarette substitute

130 assigned ENDS+8 mg/ml

120 assessed at w4

10 withdrew

164 excluded

90 screen failed at in-person screening

50 eligible but no show for randomization

24 screen failed at randomization

130 assigned ENDS+0 mg/ml

114 assessed at w12

6 withdrew

91 assessed at w24

23 withdrew

130 assigned ENDS+36 mg/ml

113 assessed at w4

17 withdrew

92 assessed at w12

21 withdrew

74 assessed at w24

18 withdrew

118 assessed at w4

12 withdrew

111 assessed at w12

7 withdrew

81 assessed at w24

30 withdrew

120 assessed at w4

10 withdrew

109 assessed at w12

11 withdrew

86 assessed at w24

23 withdrew

Supplemental table 1. Study product use percentages using intent-to-treat method (with assumption of no use to missing study product usage logs)

| **Percent using study product,**  **n (%)** | **CS** | **E-cigarette conditions** | | |
| --- | --- | --- | --- | --- |
|  |  | **0 mg/mL** | **8 mg/mL** | **36 mg/mL** |
| Week 1 | 113 (86.9) | 111 (85.4) | 114 (87.7) | 116 (89.2) |
| Week 2 | 101 (77.7) | 98 (75.4) | 110 (84.6) | 110 (84.6) |
| Week 4 | 93 (71.5) | 87 (66.9) | 98 (75.4) | 106 (81.5) |
| Week 8 | 58 (44.6) | 71 (54.6) | 74 (56.9) | 79 (60.8)* |
| Week 12 | 54 (41.5) | 55 (42.3) | 68 (52.3) | 72 (55.4)* |
| Week 16 | 45 (34.6) | 56 (43.1) | 57 (43.9) | 70 (53.9)* |
| Week 20 | 47 (36.2) | 46 (35.4) | 53 (40.8) | 63 (48.5) |
| Week 24 | 43 (33.1) | 47 (36.2) | 49 (37.7) | 62 (47.7)* |

Note: CS, cigarette substitute. Denominator for calculating the percent using is always 130 for every condition.

*p<0.05 from chi-square tests; Reference group: Cigarette substitute

Self-reported number of times product used per day (each “time” defined approximately as a period of use lasting about 15 inhalations or 10 minutes)^1^ were calculated using a 7-day timeline follow-back procedure.

^1^Foulds, J. *et al.* Development of a Questionnaire for Assessing Dependence on Electronic Cigarettes Among a Large Sample of Ex-Smoking E-cigarette Users. *Nicotine Tob. Res.* 17, 186–192 (2015).

Adapted from: Cobb CO, Foulds J, Yen MS, et al. Effect of an electronic nicotine delivery system with 0, 8, or 36 mg/mL liquid nicotine versus a cigarette substitute on tobacco-related toxicant exposure: a four-arm, parallel-group, randomised, controlled trial. Lancet Respir Med. 2021;9(8):840-850. doi:10.1016/S2213-2600(21)00022-9 (Supplementary Materials)

Supplemental table 2. Study-related adverse events by randomized groups

| **Adverse Events** | **Total** | **Cigarette substitute** | **E-cigarettes with 0 mg/mL nicotine** | **E-cigarettes with 8 mg/mL nicotine** | **E-cigarettes with 36 mg/mL nicotine** |
| --- | --- | --- | --- | --- | --- |
| **Yes, n (%)** | **N=520** | **n=130** | **n=130** | **n=130** | **n=130** |
| Abdominal pain | 1 (0.2) | 0 (0) | 1 (0.8) | 0 (0) | 0 (0) |
| Allergic reaction | 1 (0.2) | 0 (0) | 1 (0.8) | 0 (0) | 0 (0) |
| Anxiety | 4 (0.8) | 1 (0.8) | 1 (0.8) | 1 (0.8) | 1 (0.8) |
| Bloating | 1 (0.2) | 0 (0) | 0 (0) | 0 (0) | 1 (0.8) |
| Bronchial infection | 1 (0.2) | 0 (0) | 0 (0) | 1 (0.8) | 0 (0) |
| Burn | 1 (0.2) | 0 (0) | 0 (0) | 1 (0.8) | 0 (0) |
| Chest pain, cardiac | 1 (0.2) | 0 (0) | 0 (0) | 1 (0.8) | 0 (0) |
| Cough | 33 (6.3) | 0 (0) | 4 (3.1) | 18 (13.8) | 11 (8.5) |
| Depressed level of consciousness | 1 (0.2) | 0 (0) | 0 (0) | 0 (0) | 1 (0.8) |
| Depression | 1 (0.2) | 1 (0.8) | 0 (0) | 0 (0) | 0 (0) |
| Dizziness | 5 (1) | 0 (0) | 2 (1.5) | 2 (1.5) | 1 (0.8) |
| Dry mouth | 6 (1.2) | 0 (0) | 1 (0.8) | 2 (1.5) | 3 (2.3) |
| Dyspnea | 1 (0.2) | 0 (0) | 0 (0) | 1 (0.8) | 0 (0) |
| Epistaxis | 1 (0.2) | 0 (0) | 0 (0) | 0 (0) | 1 (0.8) |
| Fatigue | 2 (0.4) | 1 (0.8) | 0 (0) | 1 (0.8) | 0 (0) |
| Flu-like symptoms | 1 (0.2) | 0 (0) | 1 (0.8) | 0 (0) | 0 (0) |
| Gastrointestinal disorders, other specify | 1 (0.2) | 0 (0) | 0 (0) | 0 (0) | 1 (0.8) |
| Headache | 21 (4) | 0 (0) | 7 (5.4) | 7 (5.4) | 7 (5.4) |
| Hiccups | 2 (0.4) | 0 (0) | 0 (0) | 0 (0) | 2 (1.5) |
| Hypertension | 1 (0.2) | 0 (0) | 1 (0.8) | 0 (0) | 0 (0) |
| Injury, poisoning and procedural complications | 1 (0.2) | 0 (0) | 0 (0) | 1 (0.8) | 0 (0) |
| Irritability | 2 (0.4) | 1 (0.8) | 0 (0) | 1 (0.8) | 0 (0) |
| Mouth ulcers | 12 (2.3) | 0 (0) | 2 (1.5) | 5 (3.8) | 5 (3.8) |
| Mucus in throat sinus | 2 (0.4) | 0 (0) | 2 (1.5) | 0 (0) | 0 (0) |
| Nausea | 11 (2.1) | 0 (0) | 2 (1.5) | 4 (3.1) | 5 (3.8) |
| Non-MedDRA bad taste resulting from dry puff | 1 (0.2) | 0 (0) | 0 (0) | 1 (0.8) | 0 (0) |
| Oral pain | 2 (0.4) | 0 (0) | 1 (0.8) | 1 (0.8) | 0 (0) |
| Other metabolism nutrition disorder | 1 (0.2) | 0 (0) | 0 (0) | 1 (0.8) | 0 (0) |
| Psychiatric disorders, other specify | 2 (0.4) | 0 (0) | 0 (0) | 1 (0.8) | 1 (0.8) |
| Other respiratory symptoms | 6 (1.2) | 0 (0) | 0 (0) | 0 (0) | 6 (4.6) |
| Shortness of breath | 2 (0.4) | 0 (0) | 1 (0.8) | 1 (0.8) | 0 (0) |
| Sinus pain | 1 (0.2) | 0 (0) | 1 (0.8) | 0 (0) | 0 (0) |
| Sinus tachycardia | 1 (0.2) | 0 (0) | 0 (0) | 1 (0.8) | 0 (0) |
| Skin and subcutaneous tissue disorders, other specify | 1 (0.2) | 0 (0) | 0 (0) | 1 (0.8) | 0 (0) |
| Sore throat | 21 (4) | 0 (0) | 6 (4.6) | 5 (3.8) | 10 (7.7) |
| Throat irritation | 4 (0.8) | 0 (0) | 1 (0.8) | 1 (0.8) | 2 (1.5) |
| Upper respiratory infection | 3 (0.6) | 0 (0) | 2 (1.5) | 1 (0.8) | 0 (0) |
| Watering eyes | 1 (0.2) | 0 (0) | 0 (0) | 0 (0) | 1 (0.8) |

Note: Study-related adverse events include those rated as possibly, probably or definitely related to study product or study procedures.

Supplemental table 3. Adverse Events by randomized groups

| **Adverse Events** | **Total** | **Cigarette substitute** | **E-cigarettes with 0 mg/mL nicotine** | **E-cigarettes with 8 mg/mL nicotine** | **E-cigarettes with 36 mg/mL nicotine** |
| --- | --- | --- | --- | --- | --- |
| **Yes, n (%)** | **N=520** | **n=130** | **n=130** | **n=130** | **n=130** |
| Abdominal pain | 7 (1.3) | 1 (0.8) | 3 (2.3) | 2 (1.5) | 1 (0.8) |
| Adrenal insufficiency | 1 (0.2) | 0 (0) | 0 (0) | 0 (0) | 1 (0.8) |
| Allergic reaction | 6 (1.2) | 1 (0.8) | 1 (0.8) | 1 (0.8) | 3 (2.3) |
| Allergic rhinitis | 7 (1.3) | 0 (0) | 2 (1.5) | 2 (1.5) | 3 (2.3) |
| Anaphylaxis | 1 (0.2) | 1 (0.8) | 0 (0) | 0 (0) | 0 (0) |
| Ankle fracture | 1 (0.2) | 0 (0) | 1 (0.8) | 0 (0) | 0 (0) |
| Anxiety | 15 (2.9) | 2 (1.5) | 4 (3.1) | 5 (3.8) | 4 (3.1) |
| Appendicitis | 1 (0.2) | 0 (0) | 0 (0) | 1 (0.8) | 0 (0) |
| Appendicitis perforated | 2 (0.4) | 0 (0) | 0 (0) | 1 (0.8) | 1 (0.8) |
| Arthritis | 5 (1) | 3 (2.3) | 1 (0.8) | 1 (0.8) | 0 (0) |
| Aspiration | 3 (0.6) | 2 (1.5) | 0 (0) | 0 (0) | 1 (0.8) |
| Back pain | 18 (3.5) | 4 (3.1) | 2 (1.5) | 4 (3.1) | 8 (6.2) |
| Bladder infection | 2 (0.4) | 1 (0.8) | 0 (0) | 1 (0.8) | 0 (0) |
| Bloating | 1 (0.2) | 0 (0) | 0 (0) | 0 (0) | 1 (0.8) |
| Blurred vision | 1 (0.2) | 0 (0) | 0 (0) | 0 (0) | 1 (0.8) |
| Bone pain | 1 (0.2) | 0 (0) | 1 (0.8) | 0 (0) | 0 (0) |
| Bronchial infection | 9 (1.7) | 2 (1.5) | 3 (2.3) | 4 (3.1) | 0 (0) |
| Bruising | 2 (0.4) | 2 (1.5) | 0 (0) | 0 (0) | 0 (0) |
| Burn | 1 (0.2) | 0 (0) | 0 (0) | 1 (0.8) | 0 (0) |
| Cardiac disorders, other specify | 2 (0.4) | 0 (0) | 1 (0.8) | 0 (0) | 1 (0.8) |
| Chest pain, cardiac | 6 (1.2) | 2 (1.5) | 1 (0.8) | 1 (0.8) | 2 (1.5) |
| Chest wall pain | 2 (0.4) | 0 (0) | 0 (0) | 1 (0.8) | 1 (0.8) |
| Cholesterol high | 2 (0.4) | 1 (0.8) | 0 (0) | 1 (0.8) | 0 (0) |
| Colonic obstruction | 1 (0.2) | 0 (0) | 0 (0) | 0 (0) | 1 (0.8) |
| Conjunctivitis | 1 (0.2) | 0 (0) | 0 (0) | 0 (0) | 1 (0.8) |
| Constipation | 3 (0.6) | 1 (0.8) | 0 (0) | 1 (0.8) | 1 (0.8) |
| Cough | 48 (9.2) | 3 (2.3) | 9 (6.9) | 22 (16.9) | 14 (10.8) |
| Dehydration | 2 (0.4) | 1 (0.8) | 0 (0) | 1 (0.8) | 0 (0) |
| Depressed level of consciousness | 1 (0.2) | 0 (0) | 0 (0) | 0 (0) | 1 (0.8) |
| Depression | 20 (3.8) | 5 (3.8) | 6 (4.6) | 5 (3.8) | 4 (3.1) |
| Diarrhea | 2 (0.4) | 0 (0) | 1 (0.8) | 1 (0.8) | 0 (0) |
| Dizziness | 7 (1.3) | 1 (0.8) | 3 (2.3) | 2 (1.5) | 1 (0.8) |
| Dry mouth | 6 (1.2) | 0 (0) | 1 (0.8) | 2 (1.5) | 3 (2.3) |
| Dyspnea | 4 (0.8) | 0 (0) | 2 (1.5) | 1 (0.8) | 1 (0.8) |
| Ear and labyrinth disorders, ear infection | 1 (0.2) | 0 (0) | 1 (0.8) | 0 (0) | 0 (0) |
| Edema limbs | 1 (0.2) | 0 (0) | 0 (0) | 0 (0) | 1 (0.8) |
| Epistaxis | 2 (0.4) | 0 (0) | 0 (0) | 1 (0.8) | 1 (0.8) |
| Eye disorders, other specify | 1 (0.2) | 0 (0) | 0 (0) | 1 (0.8) | 0 (0) |
| Fall | 4 (0.8) | 1 (0.8) | 1 (0.8) | 2 (1.5) | 0 (0) |
| Fatigue | 3 (0.6) | 1 (0.8) | 0 (0) | 2 (1.5) | 0 (0) |
| Fever | 3 (0.6) | 0 (0) | 0 (0) | 1 (0.8) | 2 (1.5) |
| Flu-like symptoms | 28 (5.4) | 6 (4.6) | 7 (5.4) | 8 (6.2) | 7 (5.4) |
| Fracture | 2 (0.4) | 0 (0) | 1 (0.8) | 0 (0) | 1 (0.8) |
| Gastritis | 1 (0.2) | 0 (0) | 0 (0) | 0 (0) | 1 (0.8) |
| Gastroesophageal reflux disease | 2 (0.4) | 0 (0) | 0 (0) | 0 (0) | 2 (1.5) |
| Gastrointestinal disorders, other specify | 8 (1.5) | 2 (1.5) | 0 (0) | 2 (1.5) | 4 (3.1) |
| Gastrointestinal pain | 2 (0.4) | 0 (0) | 0 (0) | 2 (1.5) | 0 (0) |
| General disorders and administration site conditions, other specify | 2 (0.4) | 0 (0) | 1 (0.8) | 1 (0.8) | 0 (0) |
| Generalized muscle weakness | 1 (0.2) | 0 (0) | 0 (0) | 0 (0) | 1 (0.8) |
| Gum infection | 1 (0.2) | 0 (0) | 1 (0.8) | 0 (0) | 0 (0) |
| Hallucinations | 1 (0.2) | 0 (0) | 1 (0.8) | 0 (0) | 0 (0) |
| Hepatobiliary disorders | 1 (0.2) | 0 (0) | 0 (0) | 0 (0) | 1 (0.8) |
| Headache | 30 (5.8) | 2 (1.5) | 8 (6.2) | 10 (7.7) | 10 (7.7) |
| Hiccups | 2 (0.4) | 0 (0) | 0 (0) | 0 (0) | 2 (1.5) |
| Hypertension | 29 (5.6) | 5 (3.8) | 7 (5.4) | 8 (6.2) | 9 (6.9) |
| Hypertriglyceridemia | 2 (0.4) | 0 (0) | 0 (0) | 1 (0.8) | 1 (0.8) |
| Hypocalcemia | 1 (0.2) | 0 (0) | 0 (0) | 0 (0) | 1 (0.8) |
| Hypoglycemia | 1 (0.2) | 1 (0.8) | 0 (0) | 0 (0) | 0 (0) |
| Increased appetite | 1 (0.2) | 0 (0) | 0 (0) | 1 (0.8) | 0 (0) |
| Infections and infestations | 6 (1.2) | 1 (0.8) | 1 (0.8) | 1 (0.8) | 3 (2.3) |
| Injury, poisoning and procedural complications | 18 (3.5) | 5 (3.8) | 2 (1.5) | 6 (4.6) | 5 (3.8) |
| Irritability | 6 (1.2) | 2 (1.5) | 0 (0) | 2 (1.5) | 2 (1.5) |
| Investigations, other specify | 2 (0.4) | 0 (0) | 2 (1.5) | 0 (0) | 0 (0) |
| Joint range of motion decreased | 2 (0.4) | 1 (0.8) | 1 (0.8) | 0 (0) | 0 (0) |
| Laryngitis | 2 (0.4) | 1 (0.8) | 0 (0) | 0 (0) | 1 (0.8) |
| Lethargy | 1 (0.2) | 0 (0) | 0 (0) | 1 (0.8) | 0 (0) |
| Lymph node pain | 1 (0.2) | 0 (0) | 0 (0) | 0 (0) | 1 (0.8) |
| Metabolism and nutrition disorders, other specify | 1 (0.2) | 0 (0) | 0 (0) | 0 (0) | 1 (0.8) |
| Musculoskeletal and connective tissue disorder, other specify | 6 (1.2) | 1 (0.8) | 0 (0) | 4 (3.1) | 1 (0.8) |
| Musculoskeletal deformity | 1 (0.2) | 0 (0) | 1 (0.8) | 0 (0) | 0 (0) |
| Mouth ulcers | 13 (2.5) | 0 (0) | 3 (2.3) | 5 (3.8) | 5 (3.8) |
| Mucus in throat sinus | 4 (0.8) | 0 (0) | 3 (2.3) | 1 (0.8) | 0 (0) |
| Nasal congestion | 26 (5) | 6 (4.6) | 5 (3.8) | 7 (5.4) | 8 (6.2) |
| Nausea | 17 (3.3) | 2 (1.5) | 2 (1.5) | 7 (5.4) | 6 (4.6) |
| Neck pain | 1 (0.2) | 0 (0) | 0 (0) | 0 (0) | 1 (0.8) |
| Neoplasms, benign/malignant and unspecified including cysts and polyps, other specify | 4 (0.8) | 1 (0.8) | 1 (0.8) | 1 (0.8) | 1 (0.8) |
| Non-cardiac chest pain | 1 (0.2) | 1 (0.8) | 0 (0) | 0 (0) | 0 (0) |
| Non-MedDRA bad taste resulting from dry puff | 1 (0.2) | 0 (0) | 0 (0) | 1 (0.8) | 0 (0) |
| Oral pain | 4 (0.8) | 1 (0.8) | 1 (0.8) | 2 (1.5) | 0 (0) |
| Other metabolism nutrition disorder | 1 (0.2) | 0 (0) | 0 (0) | 1 (0.8) | 0 (0) |
| Pain | 17 (3.3) | 6 (4.6) | 2 (1.5) | 3 (2.3) | 6 (4.6) |
| Pain in extremity | 5 (1) | 2 (1.5) | 1 (0.8) | 2 (1.5) | 0 (0) |
| Palpitations | 1 (0.2) | 0 (0) | 1 (0.8) | 0 (0) | 0 (0) |
| Paresthesia | 1 (0.2) | 0 (0) | 0 (0) | 0 (0) | 1 (0.8) |
| Pharyngitis | 3 (0.6) | 2 (1.5) | 0 (0) | 1 (0.8) | 0 (0) |
| Pneumonitis | 3 (0.6) | 1 (0.8) | 0 (0) | 0 (0) | 2 (1.5) |
| Productive cough | 1 (0.2) | 0 (0) | 0 (0) | 1 (0.8) | 0 (0) |
| Psychiatric disorders, other specify | 7 (1.3) | 2 (1.5) | 0 (0) | 3 (2.3) | 2 (1.5) |
| Rash | 2 (0.4) | 0 (0) | 0 (0) | 2 (1.5) | 0 (0) |
| Rectal obstruction | 1 (0.2) | 0 (0) | 1 (0.8) | 0 (0) | 0 (0) |
| Reproductive system and breast disorders, other specify | 4 (0.8) | 1 (0.8) | 0 (0) | 1 (0.8) | 2 (1.5) |
| Other respiratory symptoms | 75 (14.4) | 21 (16.2) | 15 (11.5) | 14 (10.8) | 25 (19.2) |
| Rhinitis | 1 (0.2) | 0 (0) | 0 (0) | 0 (0) | 1 (0.8) |
| Shortness of breath | 3 (0.6) | 1 (0.8) | 1 (0.8) | 1 (0.8) | 0 (0) |
| Sinusitis | 12 (2.3) | 3 (2.3) | 3 (2.3) | 5 (3.8) | 1 (0.8) |
| Sinus disorder | 1 (0.2) | 0 (0) | 1 (0.8) | 0 (0) | 0 (0) |
| Skin infection | 1 (0.2) | 0 (0) | 0 (0) | 0 (0) | 1 (0.8) |
| Sinus pain | 2 (0.4) | 0 (0) | 1 (0.8) | 1 (0.8) | 0 (0) |
| Sinus tachycardia | 2 (0.4) | 0 (0) | 1 (0.8) | 1 (0.8) | 0 (0) |
| Skin and subcutaneous tissue disorders, other specify | 6 (1.2) | 1 (0.8) | 2 (1.5) | 2 (1.5) | 1 (0.8) |
| Stomach pain | 2 (0.4) | 1 (0.8) | 1 (0.8) | 0 (0) | 0 (0) |
| Sore throat | 23 (4.4) | 0 (0) | 7 (5.4) | 6 (4.6) | 10 (7.7) |
| Stroke | 1 (0.2) | 0 (0) | 0 (0) | 1 (0.8) | 0 (0) |
| Suicide attempt | 1 (0.2) | 0 (0) | 1 (0.8) | 0 (0) | 0 (0) |
| Suicidal ideation | 1 (0.2) | 0 (0) | 1 (0.8) | 0 (0) | 0 (0) |
| Surgical and medical procedures, other specify | 37 (7.1) | 7 (5.4) | 10 (7.7) | 7 (5.4) | 13 (10) |
| Syncope | 1 (0.2) | 0 (0) | 1 (0.8) | 0 (0) | 0 (0) |
| Toothache | 8 (1.5) | 2 (1.5) | 3 (2.3) | 2 (1.5) | 1 (0.8) |
| Tooth infection | 5 (1) | 1 (0.8) | 4 (3.1) | 0 (0) | 0 (0) |
| Throat irritation | 4 (0.8) | 0 (0) | 1 (0.8) | 1 (0.8) | 2 (1.5) |
| Upper respiratory infection | 37 (7.1) | 4 (3.1) | 10 (7.7) | 12 (9.2) | 11 (8.5) |
| Urinary tract infection | 3 (0.6) | 1 (0.8) | 0 (0) | 1 (0.8) | 1 (0.8) |
| Vomiting | 3 (0.6) | 1 (0.8) | 1 (0.8) | 0 (0) | 1 (0.8) |
| Watering eyes | 2 (0.4) | 0 (0) | 1 (0.8) | 0 (0) | 1 (0.8) |
| Wound infection | 1 (0.2) | 0 (0) | 0 (0) | 0 (0) | 1 (0.8) |
| Wrist fracture | 1 (0.2) | 0 (0) | 0 (0) | 0 (0) | 1 (0.8) |

Supplemental table 4. ****Study-related adverse events**** reported in the first month by randomized groups

| **Adverse Events** | **Total** | **Cigarette substitute** | **E-cigarettes with 0 mg/mL nicotine** | **E-cigarettes with 8 mg/mL nicotine** | **E-cigarettes with 36 mg/mL nicotine** |
| --- | --- | --- | --- | --- | --- |
| **Yes, n (%)** | **N=520** | **n=130** | **n=130** | **n=130** | **n=130** |
| Abdominal pain | 1 (0.2) | 0 (0) | 1 (0.8) | 0 (0) | 0 (0) |
| Allergic reaction | 1 (0.2) | 0 (0) | 1 (0.8) | 0 (0) | 0 (0) |
| Anxiety | 3 (0.6) | 0 (0) | 1 (0.8) | 1 (0.8) | 1 (0.8) |
| Burn | 1 (0.2) | 0 (0) | 0 (0) | 1 (0.8) | 0 (0) |
| Chest pain, cardiac | 1 (0.2) | 0 (0) | 0 (0) | 1 (0.8) | 0 (0) |
| Cough | 22 (4.2) | 0 (0) | 3 (2.3) | 11 (8.5) | 8 (6.2) |
| Depressed level of consciousness | 1 (0.2) | 0 (0) | 0 (0) | 0 (0) | 1 (0.8) |
| Depression | 1 (0.2) | 1 (0.8) | 0 (0) | 0 (0) | 0 (0) |
| Dizziness | 5 (1) | 0 (0) | 2 (1.5) | 2 (1.5) | 1 (0.8) |
| Dry mouth | 4 (0.8) | 0 (0) | 1 (0.8) | 0 (0) | 3 (2.3) |
| Dyspnea | 1 (0.2) | 0 (0) | 0 (0) | 1 (0.8) | 0 (0) |
| Fatigue | 1 (0.2) | 1 (0.8) | 0 (0) | 0 (0) | 0 (0) |
| Flu-like symptoms | 1 (0.2) | 0 (0) | 1 (0.8) | 0 (0) | 0 (0) |
| Gastrointestinal disorders, other specify | 1 (0.2) | 0 (0) | 0 (0) | 0 (0) | 1 (0.8) |
| Headache | 15 (2.9) | 0 (0) | 7 (5.4) | 5 (3.8) | 3 (2.3) |
| Hiccups | 1 (0.2) | 0 (0) | 0 (0) | 0 (0) | 1 (0.8) |
| Irritability | 2 (0.4) | 1 (0.8) | 0 (0) | 1 (0.8) | 0 (0) |
| Mouth ulcers | 10 (1.9) | 0 (0) | 2 (1.5) | 4 (3.1) | 4 (3.1) |
| Mucus in throat sinus | 2 (0.4) | 0 (0) | 2 (1.5) | 0 (0) | 0 (0) |
| Nausea | 7 (1.3) | 0 (0) | 1 (0.8) | 2 (1.5) | 4 (3.1) |
| Non-MedDRA bad taste resulting from dry puff | 1 (0.2) | 0 (0) | 0 (0) | 1 (0.8) | 0 (0) |
| Other metabolism nutrition disorder | 1 (0.2) | 0 (0) | 0 (0) | 1 (0.8) | 0 (0) |
| Psychiatric disorders, other specify | 1 (0.2) | 0 (0) | 0 (0) | 1 (0.8) | 0 (0) |
| Other respiratory symptoms | 6 (1.2) | 0 (0) | 0 (0) | 0 (0) | 6 (4.6) |
| Shortness of breath | 1 (0.2) | 0 (0) | 1 (0.8) | 0 (0) | 0 (0) |
| Sinus tachycardia | 1 (0.2) | 0 (0) | 0 (0) | 1 (0.8) | 0 (0) |
| Skin and subcutaneous tissue disorders, other specify | 1 (0.2) | 0 (0) | 0 (0) | 1 (0.8) | 0 (0) |
| Sore throat | 14 (2.7) | 0 (0) | 4 (3.1) | 4 (3.1) | 6 (4.6) |
| Throat irritation | 3 (0.6) | 0 (0) | 0 (0) | 1 (0.8) | 2 (1.5) |
| Upper respiratory infection | 3 (0.6) | 0 (0) | 2 (1.5) | 1 (0.8) | 0 (0) |
| Watering eyes | 1 (0.2) | 0 (0) | 0 (0) | 0 (0) | 1 (0.8) |

Note: Study-related adverse events include those rated as possibly, probably or definitely related to study product or study procedures.

Supplemental table 5. Adverse Events reported in the first month by randomized groups

| **Adverse Events** | **Total** | **Cigarette substitute** | **E-cigarettes with 0 mg/mL nicotine** | **E-cigarettes with 8 mg/mL nicotine** | **E-cigarettes with 36 mg/mL nicotine** |
| --- | --- | --- | --- | --- | --- |
| **Yes, n (%)** | **N=520** | **n=130** | **n=130** | **n=130** | **n=130** |
| Abdominal pain | 3 (0.6) | 0 (0) | 2 (1.5) | 1 (0.8) | 0 (0) |
| Allergic reaction | 4 (0.8) | 1 (0.8) | 1 (0.8) | 1 (0.8) | 1 (0.8) |
| Allergic rhinitis | 5 (1) | 0 (0) | 2 (1.5) | 2 (1.5) | 1 (0.8) |
| Anxiety | 8 (1.5) | 0 (0) | 2 (1.5) | 3 (2.3) | 3 (2.3) |
| Back pain | 9 (1.7) | 2 (1.5) | 1 (0.8) | 2 (1.5) | 4 (3.1) |
| Blurred vision | 1 (0.2) | 0 (0) | 0 (0) | 0 (0) | 1 (0.8) |
| Bone pain | 1 (0.2) | 0 (0) | 1 (0.8) | 0 (0) | 0 (0) |
| Bronchial infection | 3 (0.6) | 0 (0) | 2 (1.5) | 1 (0.8) | 0 (0) |
| Burn | 1 (0.2) | 0 (0) | 0 (0) | 1 (0.8) | 0 (0) |
| Cardiac disorders, other specify | 1 (0.2) | 0 (0) | 1 (0.8) | 0 (0) | 0 (0) |
| Chest pain, cardiac | 3 (0.6) | 0 (0) | 1 (0.8) | 1 (0.8) | 1 (0.8) |
| Chest wall pain | 2 (0.4) | 0 (0) | 0 (0) | 1 (0.8) | 1 (0.8) |
| Conjunctivitis | 1 (0.2) | 0 (0) | 0 (0) | 0 (0) | 1 (0.8) |
| Constipation | 1 (0.2) | 1 (0.8) | 0 (0) | 0 (0) | 0 (0) |
| Cough | 28 (5.4) | 0 (0) | 7 (5.4) | 12 (9.2) | 9 (6.9) |
| Dehydration | 1 (0.2) | 1 (0.8) | 0 (0) | 0 (0) | 0 (0) |
| Depressed level of consciousness | 1 (0.2) | 0 (0) | 0 (0) | 0 (0) | 1 (0.8) |
| Depression | 6 (1.2) | 3 (2.3) | 2 (1.5) | 0 (0) | 1 (0.8) |
| Diarrhea | 2 (0.4) | 0 (0) | 1 (0.8) | 1 (0.8) | 0 (0) |
| Dizziness | 7 (1.3) | 1 (0.8) | 3 (2.3) | 2 (1.5) | 1 (0.8) |
| Dry mouth | 4 (0.8) | 0 (0) | 1 (0.8) | 0 (0) | 3 (2.3) |
| Dyspnea | 2 (0.4) | 0 (0) | 1 (0.8) | 1 (0.8) | 0 (0) |
| Epistaxis | 1 (0.2) | 0 (0) | 0 (0) | 1 (0.8) | 0 (0) |
| Eye disorders, other specify | 1 (0.2) | 0 (0) | 0 (0) | 1 (0.8) | 0 (0) |
| Fall | 1 (0.2) | 0 (0) | 0 (0) | 1 (0.8) | 0 (0) |
| Fatigue | 1 (0.2) | 1 (0.8) | 0 (0) | 0 (0) | 0 (0) |
| Fever | 1 (0.2) | 0 (0) | 0 (0) | 0 (0) | 1 (0.8) |
| Flu-like symptoms | 17 (3.3) | 3 (2.3) | 3 (2.3) | 6 (4.6) | 5 (3.8) |
| Gastrointestinal disorders, other specify | 6 (1.2) | 1 (0.8) | 0 (0) | 1 (0.8) | 4 (3.1) |
| Gastrointestinal pain | 2 (0.4) | 0 (0) | 0 (0) | 2 (1.5) | 0 (0) |
| General disorders and administration site conditions, other specify |  |  |  |  |  |
| Generalized muscle weakness | 1 (0.2) | 0 (0) | 0 (0) | 0 (0) | 1 (0.8) |
| Hepatobiliary disorders | 1 (0.2) | 0 (0) | 0 (0) | 0 (0) | 1 (0.8) |
| Headache | 19 (3.7) | 2 (1.5) | 7 (5.4) | 6 (4.6) | 4 (3.1) |
| Hiccups | 1 (0.2) | 0 (0) | 0 (0) | 0 (0) | 1 (0.8) |
| Hypertension | 16 (3.1) | 1 (0.8) | 5 (3.8) | 6 (4.6) | 4 (3.1) |
| Increased appetite | 1 (0.2) | 0 (0) | 0 (0) | 1 (0.8) | 0 (0) |
| Infections and infestations | 2 (0.4) | 1 (0.8) | 0 (0) | 1 (0.8) | 0 (0) |
| Injury, poisoning and procedural complications | 4 (0.8) | 1 (0.8) | 1 (0.8) | 0 (0) | 2 (1.5) |
| Irritability | 6 (1.2) | 2 (1.5) | 0 (0) | 2 (1.5) | 2 (1.5) |
| Investigations, other specify | 1 (0.2) | 0 (0) | 1 (0.8) | 0 (0) | 0 (0) |
| Joint range of motion decreased | 1 (0.2) | 1 (0.8) | 0 (0) | 0 (0) | 0 (0) |
| Lethargy | 1 (0.2) | 0 (0) | 0 (0) | 1 (0.8) | 0 (0) |
| Lymph node pain |  |  |  |  |  |
| Metabolism and nutrition disorders, other specify | 1 (0.2) | 0 (0) | 0 (0) | 0 (0) | 1 (0.8) |
| Musculoskeletal and connective tissue disorder, other specify | 1 (0.2) | 0 (0) | 0 (0) | 1 (0.8) | 0 (0) |
| Mouth ulcers | 10 (1.9) | 0 (0) | 2 (1.5) | 4 (3.1) | 4 (3.1) |
| Mucus in throat sinus | 2 (0.4) | 0 (0) | 2 (1.5) | 0 (0) | 0 (0) |
| Nasal congestion | 11 (2.1) | 3 (2.3) | 2 (1.5) | 2 (1.5) | 4 (3.1) |
| Nausea | 11 (2.1) | 2 (1.5) | 1 (0.8) | 4 (3.1) | 4 (3.1) |
| Neck pain | 1 (0.2) | 0 (0) | 0 (0) | 0 (0) | 1 (0.8) |
| Neoplasms, benign/malignant and unspecified including cysts and polyps, other specify | 1 (0.2) | 0 (0) | 0 (0) | 1 (0.8) | 0 (0) |
| Non-MedDRA bad taste resulting from dry puff | 1 (0.2) | 0 (0) | 0 (0) | 1 (0.8) | 0 (0) |
| Other metabolism nutrition disorder | 1 (0.2) | 0 (0) | 0 (0) | 1 (0.8) | 0 (0) |
| Pain | 4 (0.8) | 1 (0.8) | 0 (0) | 1 (0.8) | 2 (1.5) |
| Pain in extremity | 1 (0.2) | 0 (0) | 0 (0) | 1 (0.8) | 0 (0) |
| Pharyngitis | 1 (0.2) | 1 (0.8) | 0 (0) | 0 (0) | 0 (0) |
| Pneumonitis | 1 (0.2) | 0 (0) | 0 (0) | 0 (0) | 1 (0.8) |
| Productive cough | 1 (0.2) | 0 (0) | 0 (0) | 1 (0.8) | 0 (0) |
| Psychiatric disorders, other specify | 2 (0.4) | 0 (0) | 0 (0) | 2 (1.5) | 0 (0) |
| Rash | 2 (0.4) | 0 (0) | 0 (0) | 2 (1.5) | 0 (0) |
| Other respiratory symptoms | 41 (7.9) | 10 (7.7) | 7 (5.4) | 6 (4.6) | 18 (13.8) |
| Shortness of breath | 1 (0.2) | 0 (0) | 1 (0.8) | 0 (0) | 0 (0) |
| Sinusitis | 6 (1.2) | 1 (0.8) | 2 (1.5) | 2 (1.5) | 1 (0.8) |
| Sinus disorder | 1 (0.2) | 0 (0) | 1 (0.8) | 0 (0) | 0 (0) |
| Skin infection | 1 (0.2) | 0 (0) | 0 (0) | 0 (0) | 1 (0.8) |
| Sinus pain | 1 (0.2) | 0 (0) | 0 (0) | 1 (0.8) | 0 (0) |
| Sinus tachycardia | 2 (0.4) | 0 (0) | 1 (0.8) | 1 (0.8) | 0 (0) |
| Skin and subcutaneous tissue disorders, other specify | 3 (0.6) | 0 (0) | 1 (0.8) | 2 (1.5) | 0 (0) |
| Stomach pain | 1 (0.2) | 0 (0) | 1 (0.8) | 0 (0) | 0 (0) |
| Sore throat | 14 (2.7) | 0 (0) | 4 (3.1) | 4 (3.1) | 6 (4.6) |
| Suicide attempt | 1 (0.2) | 0 (0) | 1 (0.8) | 0 (0) | 0 (0) |
| Suicidal ideation | 1 (0.2) | 0 (0) | 1 (0.8) | 0 (0) | 0 (0) |
| Surgical and medical procedures, other specify | 7 (1.3) | 1 (0.8) | 1 (0.8) | 3 (2.3) | 2 (1.5) |
| Toothache | 3 (0.6) | 2 (1.5) | 1 (0.8) | 0 (0) | 0 (0) |
| Tooth infection | 1 (0.2) | 0 (0) | 1 (0.8) | 0 (0) | 0 (0) |
| Throat irritation | 3 (0.6) | 0 (0) | 0 (0) | 1 (0.8) | 2 (1.5) |
| Upper respiratory infection | 13 (2.5) | 1 (0.8) | 7 (5.4) | 3 (2.3) | 2 (1.5) |
| Urinary tract infection | 1 (0.2) | 1 (0.8) | 0 (0) | 0 (0) | 0 (0) |
| Vomiting | 2 (0.4) | 1 (0.8) | 0 (0) | 0 (0) | 1 (0.8) |
| Watering eyes | 1 (0.2) | 0 (0) | 0 (0) | 0 (0) | 1 (0.8) |

Supplemental table 6. Estimation of risk difference of frequently reported ****study-related adverse events**** by randomized groups

| **Adverse events** | **Randomized comparison** | **Cigarette substitute** | **Comparison group (either e-cig with 0/8/36 mg/mL)** | **p-value** | **Risk difference**  **(CI), %** |
| --- | --- | --- | --- | --- | --- |
| **Yes, n (%)** |  | **n= 120** | **n = 120, each group** |  |  |
| Cough | E-cig with 0 mg/mL vs. cig-sub | 0 (0) | 4 (3.1) | 0.1221 | 3.1  (0.7 – 6.3) |
|  | E-cig with 8 mg/mL vs. cig-sub | 0 (0) | 18 (13.8) | **<0.0001*** | 13.9  (8.3 – 19.7) |
|  | E-cig with 36 mg/mL vs. cig-sub | 0 (0) | 11 (8.5) | **0.0008*** | 8.5  (4.1 – 13.1) |
| Dizziness | E-cig with 0 mg/mL vs. cig-sub | 0 (0) | 2 (1.5) | 0.4981 | 1.5  (0 – 3.8) |
|  | E-cig with 8 mg/mL vs. cig-sub | 0 (0) | 2 (1.5) | 0.4981 | 1.5  (0 – 3.9) |
|  | E-cig with 36 mg/mL vs. cig-sub | 0 (0) | 1 (0.8) | 1 | 0.8  (0 – 2.7) |
| Dry mouth | E-cig with 0 mg/mL vs. cig-sub | 0 (0) | 1 (0.8) | 1 | 0.8  (0 – 2.4) |
|  | E-cig with 8 mg/mL vs. cig-sub | 0 (0) | 2 (1.5) | 0.4981 | 1.5  (0 – 4) |
|  | E-cig with 36 mg/mL vs. cig-sub | 0 (0) | 3 (2.3) | 0.2471 | 2.3  (0 – 5) |
| Headache | E-cig with 0 mg/mL vs. cig-sub | 0 (0) | 7 (5.4) | **0.0144*** | 5.4  (1.6 – 9.3) |
|  | E-cig with 8 mg/mL vs. cig-sub | 0 (0) | 7 (5.4) | **0.0144*** | 5.4  (1.6 – 9.4) |
|  | E-cig with 36 mg/mL vs. cig-sub | 0 (0) | 7 (5.4) | **0.0144*** | 5.4  (1.6 – 9.7) |
| Mouth ulcers | E-cig with 0 mg/mL vs. cig-sub | 0 (0) | 2 (1.5) | 0.4981 | 1.5  (0 – 3.9) |
|  | E-cig with 8 mg/mL vs. cig-sub | 0 (0) | 5 (3.8) | 0.0601 | 3.9  (0.8 – 7.4) |
|  | E-cig with 36 mg/mL vs. cig-sub | 0 (0) | 5 (3.8) | 0.0601 | 3.9  (0.8 – 7.4) |
| Nausea | E-cig with 0 mg/mL vs. cig-sub | 0 (0) | 2 (1.5) | 0.4981 | 1.5  (0 – 3.9) |
|  | E-cig with 8 mg/mL vs. cig-sub | 0 (0) | 4 (3.1) | 0.1221 | 3.1  (0.7 – 6.2) |
|  | E-cig with 36 mg/mL vs. cig-sub | 0 (0) | 5 (3.8) | 0.0601 | 3.9  (0.8 – 7.5) |
| Other respiratory symptoms | E-cig with 0 mg/mL vs. cig-sub | 0 (0) | 0(0) | - | - |
|  | E-cig with 8 mg/mL vs. cig-sub | 0 (0) | 0(0) | - | - |
|  | E-cig with 36 mg/mL vs. cig-sub | 0 (0) | 6 (4.6) | **0.0295*** | 4.6  (1.4 – 8.6) |
| Sore throat | E-cig with 0 mg/mL vs. cig-sub | 0 (0) | 6 (4.6) | **0.0295*** | 4.6  (1.5 – 8.5) |
|  | E-cig with 8 mg/mL vs. cig-sub | 0 (0) | 5 (3.8) | 0.0601 | 3.9  (0.8 – 7.5) |
|  | E-cig with 36 mg/mL vs. cig-sub | 0 (0) | 10 (7.7) | **0.0016*** | 7.7  (3.4 – 12.7) |
| Throat irritation | E-cig with 0 mg/mL vs. cig-sub | 0 (0) | 1 (0.8) | 1 | 0.8  (0 – 2.5) |
|  | E-cig with 8 mg/mL vs. cig-sub | 0 (0) | 1 (0.8) | 1 | 0.8  (0 – 2.5) |
|  | E-cig with 36 mg/mL vs. cig-sub | 0 (0) | 2 (1.5) | 0.4981 | 1.5  (0 – 4.0) |

Note: Study-related adverse events include those rated as possibly, probably or definitely related to study product or study procedures.

Reference group: Cigarette substitute

p-value reported from Fisher exact test.

*p<0.05.

Supplemental table 7. Estimation of risk difference of frequently reported adverse events: e-cigarette vs. cigarette substitute groups

| **Adverse events** | **Cigarette substitute** | **E-cigarettes with 0/8/36 mg/mL nicotine** | **p-value** | **Risk difference**  **(CI),** |
| --- | --- | --- | --- | --- |
| **Yes, n (%)** | **n= 120** | **n = 390** |  |  |
| Cough | 3 (2.3) | 45 (11.5) | **0.0008*** | 9.2 (5.6 – 13.4) |
| Dizziness | 1 (0.8) | 6 (1.5) | 0.6863 | 0.8 (-1.2 – 2.5) |
| Dry mouth | 0 (0) | 6 (1.5) | 0.3445 | 1.5 (0.5 – 3.0) |
| Headache | 2 (1.5) | 28 (7.2) | **0.0156*** | 5.6 (2.0 – 8.8) |
| Mouth ulcers | 0 (0) | 13 (3.3) | **0.0454*** | 3.3 (1.8 – 5.1) |
| Nausea | 2 (1.5) | 15 (3.8) | 0.2627 | 2.3 (-0.6 – 5.2) |
| Other respiratory symptoms | 21 (16.2) | 54 (13.8) | 0.5643 | -2.3 (-9.9 – 4.9) |
| Sore throat | 0 (0) | 23 (5.9) | **0.0021*** | 5.9 (3.6 – 8.2) |
| Throat irritation | 0 (0) | 4 (1) | 0.5765 | 1.0 (0.2 – 2.1) |

Reference group: Cigarette substitute

p-value reported from Fisher exact test.

*p<0.05

Supplemental table 8. Estimation of risk difference of frequently reported adverse events by randomized groups

| **Adverse events, yes** | **Comparison**  **group** | **Cigarette substitute** | **Comparison group (either e-cig with 0/8/36 mg/mL)** | **p-value** | **Risk difference**  **(CI),** |
| --- | --- | --- | --- | --- | --- |
| **Yes, n (%)** |  | **n= 120** | **n = 120, each group** |  |  |
| Cough | E-cig with 0 mg/mL vs. cig-sub | 3 (2.3) | 9 (6.9) | 0.1366 | 3.1  (0.7 – 6.3) |
|  | E-cig with 8 mg/mL vs. cig-sub | 3 (2.3) | 22 (16.9) | **0.0001*** | 14.6  (8.0 – 21.1) |
|  | E-cig with 36 mg/mL vs. cig-sub | 3 (2.3) | 14 (10.8) | **0.0101*** | 8.5  (2.4 – 14.5) |
| Dizziness | E-cig with 0 mg/mL vs. cig-sub | 1 (0.8) | 3 (2.3) | 0.6221 | 1.5  (-1.4 – 4.5) |
|  | E-cig with 8 mg/mL vs. cig-sub | 1 (0.8) | 2 (1.5) | 1 | 0.8  (-1.7 – 3.4) |
|  | E-cig with 36 mg/mL vs. cig-sub | 1 (0.8) | 1 (0.8) | 1 | 0  (-2.2 – 2.2) |
| Dry mouth | E-cig with 0 mg/mL vs. cig-sub | 0 (0) | 1 (0.8) | 1 | 0.8  (0 – 2.5) |
|  | E-cig with 8 mg/mL vs. cig-sub | 0 (0) | 2 (1.5) | 0.4981 | 1.5  (0 – 4) |
|  | E-cig with 36 mg/mL vs. cig-sub | 0 (0) | 3 (2.3) | 0.2471 | 2.3  (0 – 5.1) |
| Headache | E-cig with 0 mg/mL vs. cig-sub | 2 (1.5) | 8 (6.2) | 0.1026 | 4.6  (0.1 – 9.9) |
|  | E-cig with 8 mg/mL vs. cig-sub | 2 (1.5) | 10 (7.7) | **0.0343*** | 6.2  (1.2 – 11.7) |
|  | E-cig with 36 mg/mL vs. cig-sub | 2 (1.5) | 10 (7.7) | **0.0343*** | 6.2  (1.2 – 11.7) |
| Mouth ulcers | E-cig with 0 mg/mL vs. cig-sub | 0 (0) | 3 (2.3) | 0.2471 | 2.3  (0 – 4.9) |
|  | E-cig with 8 mg/mL vs. cig-sub | 0 (0) | 5 (3.8) | 0.0601 | 3.9  (0.8 – 7.6) |
|  | E-cig with 36 mg/mL vs. cig-sub | 0 (0) | 5 (3.8) | 0.0601 | 3.9  (0.8 – 7.3) |
| Nausea | E-cig with 0 mg/mL vs. cig-sub | 2 (1.5) | 2 (1.5) | 1 | 0  (-2.9 – 2.9) |
|  | E-cig with 8 mg/mL vs. cig-sub | 2 (1.5) | 7 (5.4) | 0.1720 | 3.9  (-0.2 – 8.6) |
|  | E-cig with 36 mg/mL vs. cig-sub | 2 (1.5) | 6 (4.6) | 0.2814 | 3.1  (-1.3 – 7.6) |
| Other respiratory symptoms | E-cig with 0 mg/mL vs. cig-sub | 21 (16.2) | 15 (11.5) | 0.3695 | -4.6  (-13.2 – 3.9) |
|  | E-cig with 8 mg/mL vs. cig-sub | 21 (16.2) | 14 (10.8) | 0.2755 | -5.4 (-13.5 – 3.1) |
|  | E-cig with 36 mg/mL vs. cig-sub | 21 (16.2) | 25 (19.2) | 0.6262 | 3.1  (-6.1 – 12.3) |
| Sore throat | E-cig with 0 mg/mL vs. cig-sub | 0 (0) | 7 (5.4) | **0.0144*** | 5.4  (1.6 – 9.4) |
|  | E-cig with 8 mg/mL vs. cig-sub | 0 (0) | 6 (4.6) | **0.0295** | 4.6  (1.4 – 8.3) |
|  | E-cig with 36 mg/mL vs. cig-sub | 0 (0) | 10 (7.7) | **0.0016*** | 7.7  (3.1 – 12.2) |
| Throat irritation | E-cig with 0 mg/mL vs. cig-sub | 0 (0) | 1 (0.8) | 1 | 0.8  (0 – 2.4) |
|  | E-cig with 8 mg/mL vs. cig-sub | 0 (0) | 1 (0.8) | 1 | 0.8  (0 – 2.4) |
|  | E-cig with 36 mg/mL vs. cig-sub | 0 (0) | 2 (1.5) | 0.4981 | 1.5  (0 – 4.3) |

Reference group: Cigarette substitute

p-value reported from Fisher exact test.

*p<0.05

Supplemental table 9. Estimation of risk difference of frequently reported ****study-related adverse events**** in the first month: e-cigarette vs. cigarette substitute groups

| **Adverse events** | **Cigarette substitute** | **E-cigarettes with 0/8/36 mg/mL nicotine** | **p-value** | **Risk difference**  **(CI), %** |
| --- | --- | --- | --- | --- |
| **Yes, n (%)** | **n= 120** | **n = 390** |  |  |
| Cough | 0 (0) | 22 (5.6) | **0.0021*** | 5.6 (3.5 – 8.1) |
| Dizziness | 0 (0) | 5 (1.3) | 0.3383 | 1.3 (0.3 – 2.5) |
| Dry mouth | 0 (0) | 4 (1) | 0.5765 | 1.0 (0.2 – 2.1) |
| Headache | 0 (0) | 15 (3.8) | **0.0284*** | 3.9 (2.0 – 5.7) |
| Mouth ulcers | 0 (0) | 10 (2.6) | 0.0734 | 2.6 (1.0 – 4.2) |
| Nausea | 0 (0) | 7 (1.8) | 0.2009 | 1.8 (0.5 – 3.1) |
| Other respiratory symptoms | 0 (0) | 6 (1.5) | 0.3445 | 1.5 (0.3 – 3.0) |
| Sore throat | 0 (0) | 14 (3.6) | **0.0262*** | 3.6 (2.0 – 5.5) |
| Throat irritation | 0 (0) | 3 (0.8) | 0.5768 | 0.8 (0.0 – 1.8) |

Note: Study-related adverse events include those rated as possibly, probably or definitely related to study product or study procedures.

Reference group: Cigarette substitute

p-value reported from Fisher exact test.

*p<0.05

Supplemental table 10. Estimation of risk difference of frequently reported ****study-related adverse events**** in the first month by randomized groups

| **Adverse events** | **Randomized comparison** | **Cigarette substitute** | **Comparison group (either e-cig with 0/8/36 mg/mL)** | **p-value** | **Risk difference**  **(CI), %** |
| --- | --- | --- | --- | --- | --- |
| **Yes, n (%)** |  | **n= 120** | **n = 120, each group** |  |  |
| Cough | E-cig with 0 mg/mL vs. cig-sub | 0 (0) | 3 (2.3) | 0.2471 | 2.3  (0.0 – 5.0) |
|  | E-cig with 8 mg/mL vs. cig-sub | 0 (0) | 11 (8.5) | **0.0008*** | 8.5  (4.0 – 13.7) |
|  | E-cig with 36 mg/mL vs. cig-sub | 0 (0) | 8 (6.2) | **0.007*** | 6.2  (2.5 – 11.1) |
| Dizziness | E-cig with 0 mg/mL vs. cig-sub | 0 (0) | 2 (1.5) | 0.4981 | 1.5  (0 – 3.9) |
|  | E-cig with 8 mg/mL vs. cig-sub | 0 (0) | 2 (1.5) | 0.4981 | 1.5  (0 – 3.9) |
|  | E-cig with 36 mg/mL vs. cig-sub | 0 (0) | 1 (0.8) | 1 | 0.8  (0 – 2.6) |
| Dry mouth | E-cig with 0 mg/mL vs. cig-sub | 0 (0) | 1 (0.8) | 1 | 0.8  (0 – 2.4) |
|  | E-cig with 8 mg/mL vs. cig-sub | 0 (0) | 0 (0) | - | - |
|  | E-cig with 36 mg/mL vs. cig-sub | 0 (0) | 3 (2.3) | 0.2471 | 2.3  (0 – 5.3) |
| Headache | E-cig with 0 mg/mL vs. cig-sub | 0 (0) | 7 (5.4) | **0.0144*** | 5.4  (1.6 – 9.3) |
|  | E-cig with 8 mg/mL vs. cig-sub | 0 (0) | 5 (3.8) | 0.0601 | 3.9  (0.8 – 7.7) |
|  | E-cig with 36 mg/mL vs. cig-sub | 0 (0) | 3 (2.3) | 0.2471 | 2.3  (0.0 – 5.0) |
| Mouth ulcers | E-cig with 0 mg/mL vs. cig-sub | 0 (0) | 2 (1.5) | 0.4981 | 1.5  (0 – 3.9) |
|  | E-cig with 8 mg/mL vs. cig-sub | 0 (0) | 4 (3.1) | 0.1221 | 3.1  (0.7 – 6.5) |
|  | E-cig with 36 mg/mL vs. cig-sub | 0 (0) | 4 (3.1) | 0.1221 | 3.1  (0.7 – 6.5) |
| Nausea | E-cig with 0 mg/mL vs. cig-sub | 0 (0) | 1 (0.8) | 1 | 0.8  (0 – 2.5) |
|  | E-cig with 8 mg/mL vs. cig-sub | 0 (0) | 2 (1.5) | 0.4981 | 1.5  (0.0 – 4.0) |
|  | E-cig with 36 mg/mL vs. cig-sub | 0 (0) | 4 (3.1) | 0.1221 | 3.1  (0.7 – 6.4) |
| Other respiratory symptoms | E-cig with 0 mg/mL vs. cig-sub | 0 (0) | 0(0) | - | - |
|  | E-cig with 8 mg/mL vs. cig-sub | 0 (0) | 0(0) | - | - |
|  | E-cig with 36 mg/mL vs. cig-sub | 0 (0) | 6 (4.6) | **0.0295*** | 4.6  (1.5 – 8.7) |
| Sore throat | E-cig with 0 mg/mL vs. cig-sub | 0 (0) | 4 (3.1) | 0.1221 | 3.1  (0.7 – 6.5) |
|  | E-cig with 8 mg/mL vs. cig-sub | 0 (0) | 5 (3.8) | 0.0601 | 3.9  (0.8 – 7.5) |
|  | E-cig with 36 mg/mL vs. cig-sub | 0 (0) | 6 (4.6) | **0.0295*** | 4.6  (1.5 – 8.3) |
| Throat irritation | E-cig with 0 mg/mL vs. cig-sub | 0 (0) | 0 (0) | - | - |
|  | E-cig with 8 mg/mL vs. cig-sub | 0 (0) | 1 (0.8) | 1 | 0.8  (0 – 2.5) |
|  | E-cig with 36 mg/mL vs. cig-sub | 0 (0) | 2 (1.5) | 0.4981 | 1.5  (0 – 4.0) |

Note: Study-related adverse events include those rated as possibly, probably or definitely related to study product or study procedures.

Reference group: Cigarette substitute

p-value reported from Fisher exact test.

*p<0.05

Supplemental table 11. Estimation of risk difference of frequently reported adverse events in the first month: e-cigarette vs. cigarette substitute groups

| **Adverse events** | **Cigarette substitute** | **E-cigarettes with 0/8/36 mg/mL nicotine** | **p-value** | **Risk difference**  **(CI),** |
| --- | --- | --- | --- | --- |
| **Yes, n (%)** | **n= 120** | **n = 390** |  |  |
| Cough | 0 (0) | 28 (7.2) | **0.0004*** | 7.2 (4.7 – 9.8) |
| Dizziness | 1 (0.8) | 6 (1.5) | 0.6863 | 0.8 (-1.3 – 2.6) |
| Dry mouth | 0 (0) | 4 (1.0) | 0.5765 | 1.0 (0.2 – 2.3) |
| Headache | 2 (1.5) | 17 (4.4) | 0.1806 | 2.8 (-0.3 – 5.6) |
| Mouth ulcers | 0 (0) | 10 (2.6) | 0.0734 | 2.6 (1.1 – 4.3) |
| Nausea | 2 (1.5) | 9 (2.3) | 0.7390 | 0.8 (-2.1 – 3.2) |
| Other respiratory symptoms | 10 (7.7) | 31 (7.9) | 1 | 0.3 (-5.4 – 5.4) |
| Sore throat | 0 (0) | 14 (3.6) | **0.0262*** | 3.6 (1.8 – 5.6) |
| Throat irritation | 0 (0) | 3 (0.8) | 0.5768 | 0.8 (0.0 – 1.6) |

Reference group: Cigarette substitute

p-value reported from Fisher exact test.

*p<0.05

Supplemental table 12. Estimation of risk difference of frequently reported adverse events in the first month by randomized groups

| **Adverse events** | **Randomized comparison** | **Cigarette substitute** | **Comparison group (either e-cig with 0/8/36 mg/mL)** | **p-value** | **Risk difference**  **(CI),** |
| --- | --- | --- | --- | --- | --- |
| **Yes, n (%)** |  | **n= 120** | **n = 120, each group** |  |  |
| Cough | E-cig with 0 mg/mL vs. cig-sub | 0 (0) | 7 (5.4) | **0.0144*** | 5.4  (1.6 – 9.8) |
|  | E-cig with 8 mg/mL vs. cig-sub | 0 (0) | 12 (9.2) | **0.0004*** | 9.2  (4.6 – 14.5) |
|  | E-cig with 36 mg/mL vs. cig-sub | 0 (0) | 9 (6.9) | **0.0034*** | 6.9  (3.1 – 11.6) |
| Dizziness | E-cig with 0 mg/mL vs. cig-sub | 1 (0.8) | 3 (2.3) | 0.6221 | 1.5  (-1.5 – 4.9) |
|  | E-cig with 8 mg/mL vs. cig-sub | 1 (0.8) | 2 (1.5) | 1 | 0.8  (-1.6 – 3.3) |
|  | E-cig with 36 mg/mL vs. cig-sub | 1 (0.8) | 1 (0.8) | 1 | 0.0  (-2.3 – 2.1) |
| Dry mouth | E-cig with 0 mg/mL vs. cig-sub | 0 (0) | 1 (0.8) | 1 | 0.8  (0 – 2.4) |
|  | E-cig with 8 mg/mL vs. cig-sub | 0 (0) | 2 (1.5) | 0.4981 | 1.5  (0 – 4) |
|  | E-cig with 36 mg/mL vs. cig-sub | 0 (0) | 3 (2.3) | 0.2471 | 2.3  (0 – 5) |
| Headache | E-cig with 0 mg/mL vs. cig-sub | 2 (1.5) | 7 (5.4) | 0.1720 | 3.9  (-0.2 – 8.2) |
|  | E-cig with 8 mg/mL vs. cig-sub | 2 (1.5) | 6 (4.6) | 0.2814 | 3.1  (-1.0 – 7.7) |
|  | E-cig with 36 mg/mL vs. cig-sub | 2 (1.5) | 4 (3.1) | 0.6838 | 1.5  (-1.8 – 5.5) |
| Mouth ulcers | E-cig with 0 mg/mL vs. cig-sub | 0 (0) | 2 (1.5) | 0.4981 | 1.5  (0 – 3.9) |
|  | E-cig with 8 mg/mL vs. cig-sub | 0 (0) | 4 (3.1) | 0.1221 | 3.1  (0.7 – 6.2) |
|  | E-cig with 36 mg/mL vs. cig-sub | 0 (0) | 4 (3.1) | 0.1221 | 3.1  (0.7 – 6.2) |
| Nausea | E-cig with 0 mg/mL vs. cig-sub | 2 (1.5) | 1 (0.8) | 1 | -0.8  (-3.5 – 1.6) |
|  | E-cig with 8 mg/mL vs. cig-sub | 2 (1.5) | 4 (3.1) | 0.6838 | 1.5  (-2.1 – 5.4) |
|  | E-cig with 36 mg/mL vs. cig-sub | 2 (1.5) | 4 (3.1) | 0.6838 | 1.5  (-1.9 – 5.2) |
| Other respiratory symptoms | E-cig with 0 mg/mL vs. cig-sub | 10 (7.7) | 7 (5.4) | 0.6171 | - 2.3  (-8.8 – 4.1) |
|  | E-cig with 8 mg/mL vs. cig-sub | 10 (7.7) | 6 (4.6) | 0.4399 | -3.1  (-9.1 – 2.5) |
|  | E-cig with 36 mg/mL vs. cig-sub | 10 (7.7) | 18 (13.8) | 0.1605 | 6.2  (-1.8 – 13.8) |
| Sore throat | E-cig with 0 mg/mL vs. cig-sub | 0 (0) | 4 (3.1) | 0.1221 | 3.1  (0.7 – 6.5) |
|  | E-cig with 8 mg/mL vs. cig-sub | 0 (0) | 4 (3.1) | 0.1221 | 3.1  (0.7 – 6.3) |
|  | E-cig with 36 mg/mL vs. cig-sub | 0 (0) | 6 (4.6) | **0.0295*** | 4.6  (1.5 – 8.5) |
| Throat irritation | E-cig with 0 mg/mL vs. cig-sub | 0 (0) | 0 (0) | - | - |
|  | E-cig with 8 mg/mL vs. cig-sub | 0 (0) | 1 (0.8) | 1 | 0.8  (0 – 2.5) |
|  | E-cig with 36 mg/mL vs. cig-sub | 0 (0) | 2 (1.5) | 0.4981 | 1.5  (0 – 3.9) |

Reference group: Cigarette substitute

p-value reported from Fisher exact test.

*p<0.05

Supplemental table 13. Estimation of risk difference of frequently reported ****study-related adverse events**** by e-cigarette groups

| **Adverse events** | **Randomized comparison** | **E-cig with 0 mg/mL** | **Comparison group (either e-cig with 8/36 mg/mL)** | **p-value** | **Risk difference**  **(CI), %** |
| --- | --- | --- | --- | --- | --- |
| **Yes, n (%)** |  | **n= 120** | **n = 120, each group** |  |  |
| Cough | E-cig with 8 mg/mL vs. E-cig with 0 mg/mL | 4 (3.1) | 18 (13.8) | **0.0029*** | 10.8  (4.4 – 18.2) |
|  | E-cig with 36 mg/mL vs. E-cig with 0 mg/mL | 4 (3.1) | 11 (8.5) | 0.1078 | 5.4  (0.3 – 11.1) |
| Dizziness | E-cig with 8 mg/mL vs. E-cig with 0 mg/mL | 2 (1.5) | 2 (1.5) | 1 | 0  (-3 – 3.2) |
|  | E-cig with 36 mg/mL vs. E-cig with 0 mg/mL | 2 (1.5) | 1 (0.8) | 1 | -0.8  (-3.9 – 1.8) |
| Dry mouth | E-cig with 8 mg/mL vs. E-cig with 0 mg/mL | 1 (0.8) | 2 (1.5) | 1 | 0.8  (-1.8 – 3.5) |
|  | E-cig with 36 mg/mL vs. E-cig with 0 mg/mL | 1 (0.8) | 3 (2.3) | 0.6221 | 1.5  (-1.5 – 4.7) |
| Headache | E-cig with 8 mg/mL vs. E-cig with 0 mg/mL | 7 (5.4) | 7 (5.4) | 1 | 0  (-5.5 – 5.6) |
|  | E-cig with 36 mg/mL vs. E-cig with 0 mg/mL | 7 (5.4) | 7 (5.4) | 1 | 0  (-5.3 – 5.6) |
| Mouth ulcers | E-cig with 8 mg/mL vs. E-cig with 0 mg/mL | 2 (1.5) | 5 (3.8) | 0.4467 | 2.3  (-1.2 – 6.5) |
|  | E-cig with 36 mg/mL vs. E-cig with 0 mg/mL | 2 (1.5) | 5 (3.8) | 0.4467 | 2.3  (-1.3 – 6.2) |
| Nausea | E-cig with 8 mg/mL vs. E-cig with 0 mg/mL | 2 (1.5) | 4 (3.1) | 0.6838 | 1.5  (-2 – 5.2) |
|  | E-cig with 36 mg/mL vs. E-cig with 0 mg/mL | 2 (1.5) | 5 (3.8) | 0.4467 | 2.3  (-1.4 – 6.8) |
| Other respiratory symptoms | E-cig with 8 mg/mL vs. E-cig with 0 mg/mL | 0(0) | 0(0) | - | - |
|  | E-cig with 36 mg/mL vs. E-cig with 0 mg/mL | 0(0) | 6 (4.6) | **0.0295*** | 4.6  (1.5 – 8.5) |
| Sore throat | E-cig with 8 mg/mL vs. E-cig with 0 mg/mL | 6 (4.6) | 5 (3.8) | 1 | -0.8  (-5.4 – 3.9) |
|  | E-cig with 36 mg/mL vs. E-cig with 0 mg/mL | 6 (4.6) | 10 (7.7) | 0.4399 | 3.1  (-2.6 – 8.9) |
| Throat irritation | E-cig with 8 mg/mL vs. E-cig with 0 mg/mL | 1 (0.8) | 1 (0.8) | 1 | 0  (-2.2 – 2.3) |
|  | E-cig with 36 mg/mL vs. E-cig with 0 mg/mL | 1 (0.8) | 2 (1.5) | 1 | 0.8  (-1.7 – 3.5) |

Note: Study-related adverse events include those rated as possibly, probably or definitely related to study product or study procedures.

Reference group: E-cigarette with 0 mg/mL nicotine.

p-value reported from Fisher exact test.

*p<0.05

Supplemental table 14. Estimation of risk difference of frequently reported adverse events: e-cigarette with 0 vs. 8/36 mg/mL nicotine groups

| **Adverse events** | **E-cigarette with 0 mg/mL nicotine** | **E-cigarettes with 8/36 mg/mL nicotine** | **p-value** | **Risk difference**  **(CI),** |
| --- | --- | --- | --- | --- |
| **Yes, n (%)** | **n= 120** | **n = 240** |  |  |
| Cough | 9 (6.9) | 36 (13.8) | **0.0449*** | 6.9  (0.7 – 12.7) |
| Dizziness | 3 (2.3) | 3 (1.2) | 0.4049 | -1.1  (-4.4 – 1.5) |
| Dry mouth | 1 (0.8) | 5 (1.9) | 0.6682 | 1.1  (-1.2 – 3.2) |
| Headache | 8 (6.2) | 20 (7.7) | 0.6801 | 1.5  (-3.6 – 6.7) |
| Mouth ulcers | 3 (2.3) | 10 (3.8) | 0.5569 | 1.5  (-2.1 – 4.9) |
| Nausea | 2 (1.5) | 13 (5.0) | 0.1594 | 3.5  (0.2 – 6.8) |
| Other respiratory symptoms | 15 (11.5) | 39 (15) | 0.4371 | 3.5  (-3.8 – 10.5) |
| Sore throat | 7 (5.4) | 16 (6.2) | 0.8241 | 0.8  (-4.3 – 5.5) |
| Throat irritation | 1 (0.8) | 3 (1.2) | 1 | 0.4  (-1.6 – 2.2) |

Reference group: E-cigarette with 0 mg/mL nicotine.

p-value reported from Fisher exact test.

*p<0.05

Supplemental table 15. Estimation of risk difference of frequently reported adverse events by e-cigarette groups

| **Adverse events** | **Randomized comparison** | **E-cig with 0 mg/mL** | **Comparison group (either e-cig with 8/36 mg/mL)** | **p-value** | **Risk difference**  **(CI),** |
| --- | --- | --- | --- | --- | --- |
| **Yes, n (%)** |  | **n= 120** | **n = 120, each group** |  |  |
| Cough | E-cig with 8 mg/mL vs. E-cig with 0 mg/mL | 9 (6.9) | 22 (16.9) | **0.0204*** | 10.0  (2.5 – 18.0) |
|  | E-cig with 36 mg/mL vs. E-cig with 0 mg/mL | 9 (6.9) | 14 (10.8) | 0.3828 | 3.9  (-2.6 – 10.9) |
| Dizziness | E-cig with 8 mg/mL vs. E-cig with 0 mg/mL | 3 (2.3) | 2 (1.5) | 1 | -0.8  (-4.2 – 2.4) |
|  | E-cig with 36 mg/mL vs. E-cig with 0 mg/mL | 3 (2.3) | 1 (0.8) | 0.6221 | -1.5  (-4.7 – 1.3) |
| Dry mouth | E-cig with 8 mg/mL vs. E-cig with 0 mg/mL | 1 (0.8) | 2 (1.5) | 1 | 0.8  (-1.6 – 3.4) |
|  | E-cig with 36 mg/mL vs. E-cig with 0 mg/mL | 1 (0.8) | 3 (2.3) | 0.6221 | 1.5  (-1.3 – 4.7) |
| Headache | E-cig with 8 mg/mL vs. E-cig with 0 mg/mL | 8 (6.2) | 10 (7.7) | 0.8078 | 1.5  (-4.5 – 7.2) |
|  | E-cig with 36 mg/mL vs. E-cig with 0 mg/mL | 8 (6.2) | 10 (7.7) | 0.8078 | 1.5  (-4.5 – 7.2) |
| Mouth ulcers | E-cig with 8 mg/mL vs. E-cig with 0 mg/mL | 3 (2.3) | 5 (3.8) | 0.7223 | 1.5  (-2.7 – 5.5) |
|  | E-cig with 36 mg/mL vs. E-cig with 0 mg/mL | 3 (2.3) | 5 (3.8) | 0.7223 | 1.5  (-2.7 – 5.5) |
| Nausea | E-cig with 8 mg/mL vs. E-cig with 0 mg/mL | 2 (1.5) | 7 (5.4) | 0.1720 | 3.9  (-0.4 – 8.6) |
|  | E-cig with 36 mg/mL vs. E-cig with 0 mg/mL | 2 (1.5) | 6 (4.6) | 0.2814 | 3.1  (-0.8 – 7.6) |
| Other respiratory symptoms | E-cig with 8 mg/mL vs. E-cig with 0 mg/mL | 15 (11.5) | 14 (10.8) | 1 | -0.8  (-9 – 7.3) |
|  | E-cig with 36 mg/mL vs. E-cig with 0 mg/mL | 15 (11.5) | 25 (19.2) | 0.1211 | 7.7  (-0.8 – 16.6) |
| Sore throat | E-cig with 8 mg/mL vs. E-cig with 0 mg/mL | 7 (5.4) | 6 (4.6) | 1 | -0.8  (-5.8 – 4.4) |
|  | E-cig with 36 mg/mL vs. E-cig with 0 mg/mL | 7 (5.4) | 10 (7.7) | 0.6171 | 2.3  (-3.6 – 8.2) |
| Throat irritation | E-cig with 8 mg/mL vs. E-cig with 0 mg/mL | 1 (0.8) | 1 (0.8) | 1 | 0  (-2.4 – 1.8) |
|  | E-cig with 36 mg/mL vs. E-cig with 0 mg/mL | 1 (0.8) | 2 (1.5) | 1 | 0.8  (-1.7 – 3.5) |

Reference group: E-cigarette with 0 mg/mL nicotine.

p-value reported from Fisher exact test.

*p<0.05

Supplemental table 16. Estimation of risk difference of frequently reported ****study-related adverse events**** in the first month: e-cigarette with 0 vs. 8/36 mg/mL nicotine groups

| **Adverse events** | **E-cigarette with 0 mg/mL nicotine** | **E-cigarettes with 8/36 mg/mL nicotine** | **p-value** | **Risk difference**  **(CI), %** |
| --- | --- | --- | --- | --- |
| **Yes, n (%)** | **n= 120** | **n = 240** |  |  |
| Cough | 3 (2.3) | 19 (7.3) | 0.0601 | 5.0  (1.1 – 8.9) |
| Dizziness | 2 (1.5) | 3 (1.2) | 1 | -0.4  (-3.1 – 2.0) |
| Dry mouth | 1 (0.8) | 3 (1.2) | 1 | 0.4  (-1.9 – 2.3) |
| Headache | 7 (5.4) | 8 (3.1) | 0.2741 | -2.3  (-6.9 – 1.8) |
| Mouth ulcers | 2 (1.5) | 8 (3.1) | 0.5066 | 1.5  (-1.6 – 4.5) |
| Nausea | 1 (0.8) | 6 (2.3) | 0.4323 | 1.5  (-0.9 – 3.8) |
| Other respiratory symptoms | 0 (0) | 6 (2.3) | 0.1845 | 2.3  (0.8 – 4.3) |
| Sore throat | 4 (3.1) | 10 (3.8) | 0.7817 | 0.8  (-3.2 – 4.4) |
| Throat irritation | 0 (0) | 3 (1.2) | 0.5538 | 1.1  (0.0 – 2.4) |

Note: Study-related adverse events include those rated as possibly, probably or definitely related to study product or study procedures.

Reference group: E-cigarette with 0 mg/mL nicotine.

p-value reported from Fisher exact test.

*p<0.05

Supplemental table 17. Estimation of risk difference of frequently reported ****study-related adverse events**** in the first month by e-cigarette groups

| **Adverse events** | **Randomized comparison** | **E-cig with 0 mg/mL** | **Comparison group (either e-cig with 8/36 mg/mL)** | **p-value** | **Risk difference**  **(CI), %** |
| --- | --- | --- | --- | --- | --- |
| **Yes, n (%)** |  | **n= 120** | **n = 120, each group** |  |  |
| Cough | E-cig with 8 mg/mL vs. E-cig with 0 mg/mL | 3 (2.3) | 11 (8.5%) | 0.0508 | 6.2  (0.9 – 11.7) |
|  | E-cig with 36 mg/mL vs. E-cig with 0 mg/mL | 3 (2.3) | 8 (6.2%) | 0.2165 | 3.9  (-0.7 – 9.2) |
| Dizziness | E-cig with 8 mg/mL vs. E-cig with 0 mg/mL | 2 (1.5) | 2 (1.5) | 1 | 0  (-3 – 3.2) |
|  | E-cig with 36 mg/mL vs. E-cig with 0 mg/mL | 2 (1.5) | 1 (0.8) | 1 | -0.8  (-3.4 – 1.6) |
| Dry mouth | E-cig with 8 mg/mL vs. E-cig with 0 mg/mL | 1 (0.8) | 0 (0) | 1 | -0.8  (-2.5 – 0) |
|  | E-cig with 36 mg/mL vs. E-cig with 0 mg/mL | 1 (0.8) | 3 (2.3) | 0.6221 | 1.5  (-1.4 – 4.7) |
| Headache | E-cig with 8 mg/mL vs. E-cig with 0 mg/mL | 7 (5.4) | 5 (3.8) | 0.7690 | -1.5  (-6.4 – 3.4) |
|  | E-cig with 36 mg/mL vs. E-cig with 0 mg/mL | 7 (5.4) | 3 (2.3) | 0.3341 | -3.1  (-8.0 – 1.7) |
| Mouth ulcers | E-cig with 8 mg/mL vs. E-cig with 0 mg/mL | 2 (1.5) | 4 (3.1) | 0.6838 | 1.5  (-1.9 – 5.2) |
|  | E-cig with 36 mg/mL vs. E-cig with 0 mg/mL | 2 (1.5) | 4 (3.1) | 0.6828 | 1.5  (-2.1 – 5.2) |
| Nausea | E-cig with 8 mg/mL vs. E-cig with 0 mg/mL | 1 (0.8) | 2 (1.5) | 1 | 0.8  (-1.6 – 3.4) |
|  | E-cig with 36 mg/mL vs. E-cig with 0 mg/mL | 1 (0.8) | 4 (3.1) | 0.3701 | 2.3  (-0.9 – 5.7) |
| Other respiratory symptoms | E-cig with 8 mg/mL vs. E-cig with 0 mg/mL | 0(0) | 0(0) | - | - |
|  | E-cig with 36 mg/mL vs. E-cig with 0 mg/mL | 0(0) | 6 (4.6) | **0.0295*** | 4.6  (1.5 – 8.5) |
| Sore throat | E-cig with 8 mg/mL vs. E-cig with 0 mg/mL | 4 (3.1) | 4 (3.1) | 1 | - |
|  | E-cig with 36 mg/mL vs. E-cig with 0 mg/mL | 4 (3.1) | 6 (4.6) | 0.7490 | 1.5  (-3.0 – 6.1) |
| Throat irritation | E-cig with 8 mg/mL vs. E-cig with 0 mg/mL | 0 (0) | 1 (0.8) | 1 | 0.8  (0.0 – 2.6) |
|  | E-cig with 36 mg/mL vs. E-cig with 0 mg/mL | 0 (0) | 2 (1.5) | 0.4981 | 1.5  (0 – 4.0) |

Note: Study-related adverse events include those rated as possibly, probably or definitely related to study product or study procedures.

Reference group: E-cigarette with 0 mg/mL nicotine.

p-value reported from Fisher exact test.

*p<0.05

Supplemental table 18. Estimation of risk difference of frequently reported adverse events in the first month: e-cigarette with 0 vs. 8/36 mg/mL nicotine groups

| **Adverse events,** | **E-cigarette with 0 mg/mL nicotine** | **E-cigarettes with 8/36 mg/mL nicotine** | **p-value** | **Risk difference**  **(CI),** |
| --- | --- | --- | --- | --- |
| **Yes, n (%)** | **n= 120** | **n = 240** |  |  |
| Cough | 7 (5.4) | 21 (8.1) | 0.4083 | 2.7  (-2.7 – 7.6) |
| Dizziness | 3 (2.3) | 3 (1.2) | 0.4049 | -1.1  (-4.4 – 1.5) |
| Dry mouth | 1 (0.8) | 3 (1.2) | 1 | 0.4  (-1.9 – 2.2) |
| Headache | 7 (5.4) | 10 (3.8) | 0.5994 | -1.5  (-6 – 2.9) |
| Mouth ulcers | 2 (1.5) | 8 (3.1) | 0.5066 | 1.5  (-1.6 – 4.6) |
| Nausea | 1 (0.8) | 8 (3.1) | 0.2822 | 2.3  (-0.3 – 4.7) |
| Other respiratory symptoms | 7 (5.4) | 24 (9.2) | 0.2347 | 3.9  (-1.7 – 8.9) |
| Sore throat | 4 (3.1) | 10 (3.8) | 0.7817 | 0.8  (-3.4 – 4.7) |
| Throat irritation | 0 (0) | 3 (1.2) | 0.5538 | 1.1  (0 – 2.6) |

Reference group: E-cigarette with 0 mg/mL nicotine

p-value reported from Fisher exact test.

*p<0.05

Supplemental table 19. Estimation of risk difference of frequently reported adverse events in the first month by e-cigarette arms

| **Adverse events** | **Randomized comparison** | **E-cig with 0 mg/mL** | **Comparison group (either e-cig with 8/36 mg/mL)** | **p-value** | **Risk difference**  **(CI),** |
| --- | --- | --- | --- | --- | --- |
| **Yes, n (%)** |  | **n= 120** | **n = 120, each group** |  |  |
| Cough | E-cig with 8 mg/mL vs. E-cig with 0 mg/mL | 7 (5.4) | 12 (9.2) | 0.3408 | 3.9  (-2.1 – 10.8) |
|  | E-cig with 36 mg/mL vs. E-cig with 0 mg/mL | 7 (5.4) | 9 (6.9) | 0.7973 | 1.5  (-4.3 – 6.9) |
| Dizziness | E-cig with 8 mg/mL vs. E-cig with 0 mg/mL | 3 (2.3) | 2 (1.5) | 1 | -0.8  (-4.1 – 2.4) |
|  | E-cig with 36 mg/mL vs. E-cig with 0 mg/mL | 3 (2.3) | 1 (0.8) | 0.6221 | -1.5  (-4.4 – 1.3) |
| Dry mouth | E-cig with 8 mg/mL vs. E-cig with 0 mg/mL | 1 (0.8) | 0 (0) | 1 | -0.8  (-2.5 – 0.0) |
|  | E-cig with 36 mg/mL vs. E-cig with 0 mg/mL | 1 (0.8) | 3 (2.3) | 0.6221 | 1.5  (-1.4 – 4.7) |
| Headache | E-cig with 8 mg/mL vs. E-cig with 0 mg/mL | 7 (5.4) | 6 (4.6) | 1 | -0.8  (-6.2 – 4.4) |
|  | E-cig with 36 mg/mL vs. E-cig with 0 mg/mL | 7 (5.4) | 4 (3.1) | 0.5399 | -2.3  (-7.4 – 2.6) |
| Mouth ulcers | E-cig with 8 mg/mL vs. E-cig with 0 mg/mL | 2 (1.5) | 4 (3.1) | 0.6838 | 1.5  (-2.0 – 5.3) |
|  | E-cig with 36 mg/mL vs. E-cig with 0 mg/mL | 2 (1.5) | 4 (3.1) | 0.6838 | 1.5  (-1.7 – 5.5) |
| Nausea | E-cig with 8 mg/mL vs. E-cig with 0 mg/mL | 1 (0.8) | 4 (3.1) | 0.3701 | 2.3  (-0.7 – 5.8) |
|  | E-cig with 36 mg/mL vs. E-cig with 0 mg/mL | 1 (0.8) | 4 (3.1) | 0.3701 | 2.3  (-0.7 – 5.8) |
| Other respiratory symptoms | E-cig with 8 mg/mL vs. E-cig with 0 mg/mL | 7 (5.4) | 6 (4.6) | 1 | -0.8 (-5.8 – 4.4) |
|  | E-cig with 36 mg/mL vs. E-cig with 0 mg/mL | 7 (5.4) | 18 (13.8) | **0.0336*** | 8.5  (1.5 – 15.3) |
| Sore throat | E-cig with 8 mg/mL vs. E-cig with 0 mg/mL | 4 (3.1) | 4 (3.1) | 1 | 0.0  (-4.1 – 4.1) |
|  | E-cig with 36 mg/mL vs. E-cig with 0 mg/mL | 4 (3.1) | 6 (4.6) | 0.7490 | 1.5  (-3.2 – 6.5) |
| Throat irritation | E-cig with 8 mg/mL vs. E-cig with 0 mg/mL | 0 (0) | 1 (0.8) | 1 | 0.8  (0.0 – 2.5) |
|  | E-cig with 36 mg/mL vs. E-cig with 0 mg/mL | 0 (0) | 2 (1.5) | 0.4981 | 0.8  (0.0 – 4.0) |

Reference group: E-cigarette with 0 mg/mL nicotine

p-value reported from Fisher exact test.

*p<0.05

****Supplemental**** table 20****. Study-related adverse events by flavor choice among participants randomized to e-cigarette arms****

| **Adverse Events** | **Total** | **Tobacco flavor** | **Menthol flavor** | **p-value** |
| --- | --- | --- | --- | --- |
| **Yes, n (%)** | **N=390** | **n=144** | **n=246** | **n=130** |
| Abdominal pain | 1 (0.3) | 0 (0) | 1 (0.4) | 1 |
| Allergic reaction | 1 (0.3) | 0 (0) | 1 (0.4) | 1 |
| Anxiety | 3 (0.8) | 1 (0.7) | 2 (0.8) | 1 |
| Bloating | 1 (0.3) | 1 (0.7) | 0 (0) | 0.369 |
| Bronchial infection | 1 (0.3) | 0 (0) | 1 (0.4) | 1 |
| Burn | 1 (0.3) | 0 (0) | 1 (0.4) | 1 |
| Chest pain, cardiac | 1 (0.3) | 0 (0) | 1 (0.4) | 1 |
| Cough | 33 (8.5) | 16 (11.1) | 17 (6.9) | 0.187 |
| Depressed level of consciousness | 1 (0.3) | 1 (0.7) | 0 (0) | 0.369 |
| Dizziness | 5 (1.3) | 3 (2.1) | 2 (0.8) | 0.363 |
| Dry mouth | 2 (1.4) | 4 (1.6) | 2 (1.4) | 1 |
| Dyspnea | 1 (0.3) | 0 (0) | 1 (0.4) | 1 |
| Epistaxis | 1 (0.3) | 1 (0.7) | 0 (0) | 0.369 |
| Fatigue | 1 (0.3) | 0 (0) | 1 (0.4) | 1 |
| Flu-like symptoms | 1 (0.3) | 0 (0) | 1 (0.4) | 1 |
| Gastrointestinal disorders, other specify | 1 (0.3) | 0 (0) | 1 (0.4) | 1 |
| Headache | 21 (5.4) | 5 (3.5) | 16 (6.5) | 0.249 |
| Hiccups | 2 (0.5) | 1 (0.7) | 1 (0.4) | 1 |
| Hypertension | 1 (0.3) | 0 (0) | 1 (0.4) | 1 |
| Injury, poisoning and procedural complications | 1 (0.3) | 1 (0.7) | 0 (0) | 0.369 |
| Irritability | 1 (0.3) | 0 (0) | 1 (0.4) | 1 |
| Mouth ulcers | 12 (3.1) | 7 (4.9) | 5 (2) | 0.1359 |
| Mucus in throat sinus | 2 (0.5) | 1 (0.7) | 1 (0.4) | 1 |
| Nausea | 11 (2.8) | 4 (2.8) | 7 (2.8) | 1 |
| Non-MedDRA bad taste resulting from dry puff | 1 (0.3) | 1 (0.7) | 0 (0) | 0.369 |
| Oral pain | 2 (0.5) | 0 (0) | 2 (0.8) | 0.533 |
| Other metabolism nutrition disorder | 1 (0.3) | 0 (0) | 1 (0.4) | 1 |
| Psychiatric disorders, other specify | 2 (0.5) | 2 (1.4) | 0 (0) | 0.136 |
| Other respiratory symptoms | 6 (1.5) | 3 (2.1) | 3 (1.2) | 0.674 |
| Shortness of breath | 2 (0.5) | 1 (0.7) | 1 (0.4) | 1 |
| Sinus pain | 1 (0.3) | 1 (0.7) | 0 (0) | 0.369 |
| Sinus tachycardia | 1 (0.3) | 0 (0) | 1 (0.4) | 1 |
| Skin and subcutaneous tissue disorders, other specify | 1 (0.3) | 0 (0) | 1 (0.4) | 1 |
| Sore throat | 21 (5.4) | 12 (8.3) | 9 (3.7) | 0.062 |
| Throat irritation | 4 (1) | 3 (2.1) | 1 (0.4) | 0.144 |
| Upper respiratory infection | 3 (0.8) | 0 (0) | 3 (1.2) | 0.300 |
| Watering eyes | 1 (0.3) | 1 (0.7) | 0 (0) | 0.369 |

p-values reported from Fisher exact test.

*p<0.05.
